# Safety Monitoring of mRNA Vaccines Administered During the Initial 6 Months of the U.S. COVID-19 Vaccination Program: Reports to Vaccine Adverse Events Reporting System (VAERS) and v-safe

**DOI:** 10.1101/2021.10.26.21265261

**Authors:** Hannah G. Rosenblum, Julianne M. Gee, Ruiling Liu, Paige L. Marquez, Bicheng Zhang, Penelope Strid, Winston E. Abara, Michael M. McNeil, Tanya R. Myers, Anne M. Hause, John R. Su, Bethany Baer, David Menschik, Lauri E. Markowitz, Tom T. Shimabukuro, David K. Shay

## Abstract

**Background:** In December 2020, two mRNA-based COVID-19 vaccines were authorized for use in the United States. Vaccine safety was monitored using the Vaccine Adverse Event Reporting System (VAERS), a passive surveillance system, and v-safe, an active surveillance system.

**Methods:** VAERS and v-safe data during December 14, 2020—June 14, 2021 were analyzed. VAERS reports were categorized as non-serious, serious, or death; reporting rates were calculated. Rates of reported deaths were compared to expected mortality rates by age. Proportions of v-safe participants reporting local and systemic reactions or health impacts the week following doses 1 and 2 were determined.

**Findings:** During the analytic period, 298,792,852 doses of mRNA vaccines were administered in the United States. VAERS processed 340,522 reports; 92·1% were non-serious; 6·6%, serious, non-death; and 1·3%, death. Over half of 7,914,583 v-safe participants self-reported local and systemic reactogenicity, more frequently after dose 2. Injection-site pain, fatigue, and headache were commonly reported during days 0–7 following vaccination. Reactogenicity was reported most frequently one day after vaccination; most reactions were mild. More reports of being unable to work or do normal activities occurred after dose 2 (32·1%) than dose 1 (11·9%); <1% of participants reported seeking medical care after vaccination. Rates of deaths reported to VAERS were lower than expected background rates by age group.

**Interpretation:** Safety data from >298 million doses of mRNA COVID-19 vaccine administered in the first 6 months of the U.S. vaccination program show the majority of reported adverse events were mild and short in duration.

## Introduction

In December 2020, two mRNA coronavirus disease 2019 (COVID-19) vaccines (BNT162b2; Pfizer-BioNTech and mRNA-1273; Moderna) were granted Emergency Use Authorization (EUA) by the U.S. Food and Drug Administration (FDA) as 2-dose series and recommended for use by the Advisory Committee on Immunization Practices (ACIP).^1,2^ In clinical trials, both mRNA COVID-19 vaccines showed acceptable safety profiles;^3,4^ the most frequently reported local and systemic symptoms were injection site pain, fatigue, and headache. Reactogenicity was more frequently reported following dose 2, and more common among participants aged <65 years.^3-5^

Post-authorization safety monitoring better characterizes the safety profiles of mRNA-based COVID-19 vaccines in larger and more heterogeneous populations.^6^ Phased administration of COVID-19 vaccines in the United States began with healthcare workers and long-term care facility residents and expanded to the general population by Spring 2021; however, implementation plans varied by state.^7^ The major sources of initial U.S. safety data were the Vaccine Adverse Event Reporting System (VAERS), a spontaneous, passive reporting system,^8^ and v-safe,^9^ a new active monitoring system. Reports from these systems have been issued through websites, publications, and presentations to advisory committees.^10-14^ We review VAERS and v-safe data during the first 6 months of the U.S. vaccination program, when >298 million doses of mRNA COVID-19 vaccines were administered.

## Methods

### VAERS

VAERS is a national spontaneous reporting system for detecting potential adverse events (AEs) for authorized or licensed U.S. vaccines^8^ co-administered by the Centers for Disease Control and Prevention (CDC) and FDA. VAERS accepts reports from healthcare providers, manufacturers, and the public. Reports include information about the vaccinated person, type of vaccine administered, and AEs experienced. We included all VAERS reports for mRNA vaccines submitted and processed from December 14, 2020 through June 14, 2021, inclusive of any interval from vaccination to event report.^15^ Processed reports were those quality checked and coded using the Medical Dictionary for Regulatory Activities (MedDRA) terminology.^8^ Each VAERS report may be assigned more than one MedDRA Preferred Term (PT); PTs do not necessarily indicate a medically confirmed diagnosis, and include signs and symptoms of illness and results of diagnostic tests.

Based on the Code of Federal Regulations,^16^ VAERS reports were classified as ‘serious’ if any of the following were documented: hospitalization, prolongation of hospitalization, permanent disability, life-threatening illness, congenital anomaly or birth defect, or death. Adverse events of special interest were selected for enhanced COVID-19 vaccine safety monitoring based on biologic plausibility, previous vaccine safety experience, and theoretical concerns related to COVID-19.^17^ Death certificates and autopsy reports were requested for death reports. CDC physicians reviewed VAERS reports and available death certificates for each decedent to form an impression about cause of death. Impressions were assigned to one of the following categories: one of the 15 most common *International Classification of Disease, Tenth Revision (ICD-10*) diagnostic categories reported on U.S. death certificates^18^; COVID-19-related; other (i.e., impression was not included in a pre-specified categories); or unknown/unclear if a likely cause could not be determined.

### V-safe

V-safe is a voluntary smartphone-based system that uses text messaging and secure web-based surveys to actively monitor COVID-19 vaccine safety for common local injection site and systemic reactions.^9^ V-safe participants receive text messages that link to web-based health check-in surveys following vaccination, initially daily (days 0–7), then at longer intervals post vaccination. The system resets to the initial survey frequency after entry of another dose. We analyzed survey reports from days 0–7 for reactogenicity, severity^9^ (mild, moderate, severe), and health impact (i.e., unable to perform normal daily activities, unable to work, and/or received care from a medical professional). Participants who reported receiving medical care were contacted and VAERS reports were completed, if clinically indicated.

### Data analyses

We conducted descriptive analyses of available VAERS and v-safe data from December 14, 2020 through June 14, 2021 following dose 1 and dose 2 of BNT162b2 and mRNA-1273 vaccines among individuals aged ≥16 years. For VAERS, bivariate analyses included sex, age groups, race/ethnicity, serious AEs, and vaccine type/manufacturer administered and for death reports, time from vaccination to reported death (i.e., onset interval) and cause of death. Reporting rates to VAERS were calculated for AEs using the total number of doses of mRNA vaccine administered during the 6-month period. COVID-19 vaccine administration data were provided through CDC’s COVID-19 Data Tracker.^19^ Reporting rates for deaths within 7 days following vaccination and 42 days following vaccination were calculated per vaccinated persons using persons who received at least one dose as the estimation of persons vaccinated and compared to expected background mortality rates.^20^

Empirical Bayesian (EB) data mining was used to detect disproportional reporting of post-vaccine outcomes by vaccine among all VAERS serious and non-serious reports received by June 14, 2021.^21^ This statistical method calculates observed to expected MedDRA PT pairings by comparing a specific vaccine-MedDRA PT pair to all vaccine-PT pairs in VAERS, adjusting for age, sex, and year received.^22^ These ratios are ranked by the lower 5% bound of the EB geometric mean confidence interval (EB05) and a standard alert threshold of EB05 >2 was used. An EB05 >2 represents a high degree of confidence that a vaccine-PT pair was reported at least twice as frequently as expected.

V-safe participants who responded to at least one health check-in survey during day 0–7 after vaccination were included in analyses. Descriptive statistics were calculated for participants’ characteristics (sex, age, race/ethnicity), reaction (type and severity) and health impact by manufacturer, dose number, and number of days following vaccination.

SAS software, version 9.4 (SAS Institute; Cary, NC, USA) was used for analyses. Both VAERS and v-safe conduct surveillance as a public health function and are exempt from institutional review board review. Activities were reviewed by the CDC and conducted in accordance with applicable federal law and CDC policy (See: 45 C.F.R. part 46.102(l)(2), 21 C.F.R. part 56; 42 U.S.C. §241(d); 5 U.S.C. §552a; 44 U.S.C. §3501 et seq.).

## Results

During December 14, 2020–June 14, 2021, a total of 298,792,852 doses of mRNA COVID-19 vaccines were administered in the United States: 167,177,332 were BNT162b2 and 131,639,515 were mRNA-1273 (Supplemental Table 1). A greater proportion of vaccines were administered to females (53·2%) compared with males (45·8%). The median age at vaccination was 50 years (inter-quartile range [IQR]: 33–65) for BNT162b2 and 56 years (IQR: 39–68) for mRNA-1273, respectively. Non-Hispanic White persons accounted for 38·4% of vaccine recipients; however, race/ethnicity was unknown for 34·9% of all vaccine recipients.

### VAERS

During the analytic period, VAERS received and processed a total of 340,522 reports: 164,669 following BNT162b2 and 175,816 following mRNA-1273 vaccination (Table 1). Of these reports, 92·1% were classified as non-serious, 6·6% were serious, not resulting in a death (non-death), and 1·3% were deaths. Seventy-two percent of reports were among females, and 45·3% of reports were among those aged 18–49 years; median age was 50 years (IQR: 36–64). Fifty percent of those reporting race/ethnicity identified as non-Hispanic White; for 22·1%, race/ethnicity was unknown. The most common MedDRA PTs among non-serious reports were headache (20·4%), fatigue (16·6%), pyrexia (16·3%), chills (15·7%), and pain (15·2%). The most common MedDRA PTs among serious reports were dyspnea (15·4%), death (14·1%), pyrexia (11·0%), fatigue (9·7%), and headache (9·5%). The reporting rate to VAERS was 1,049 non-serious reports per million doses, and 90 serious reports per million doses (Table 2). Among the pre-specified AESIs, reporting rates ranged from 0·1 narcolepsy reports per million doses administered to 32 COVID-19 disease reports per million doses administered.

**Table 1:**
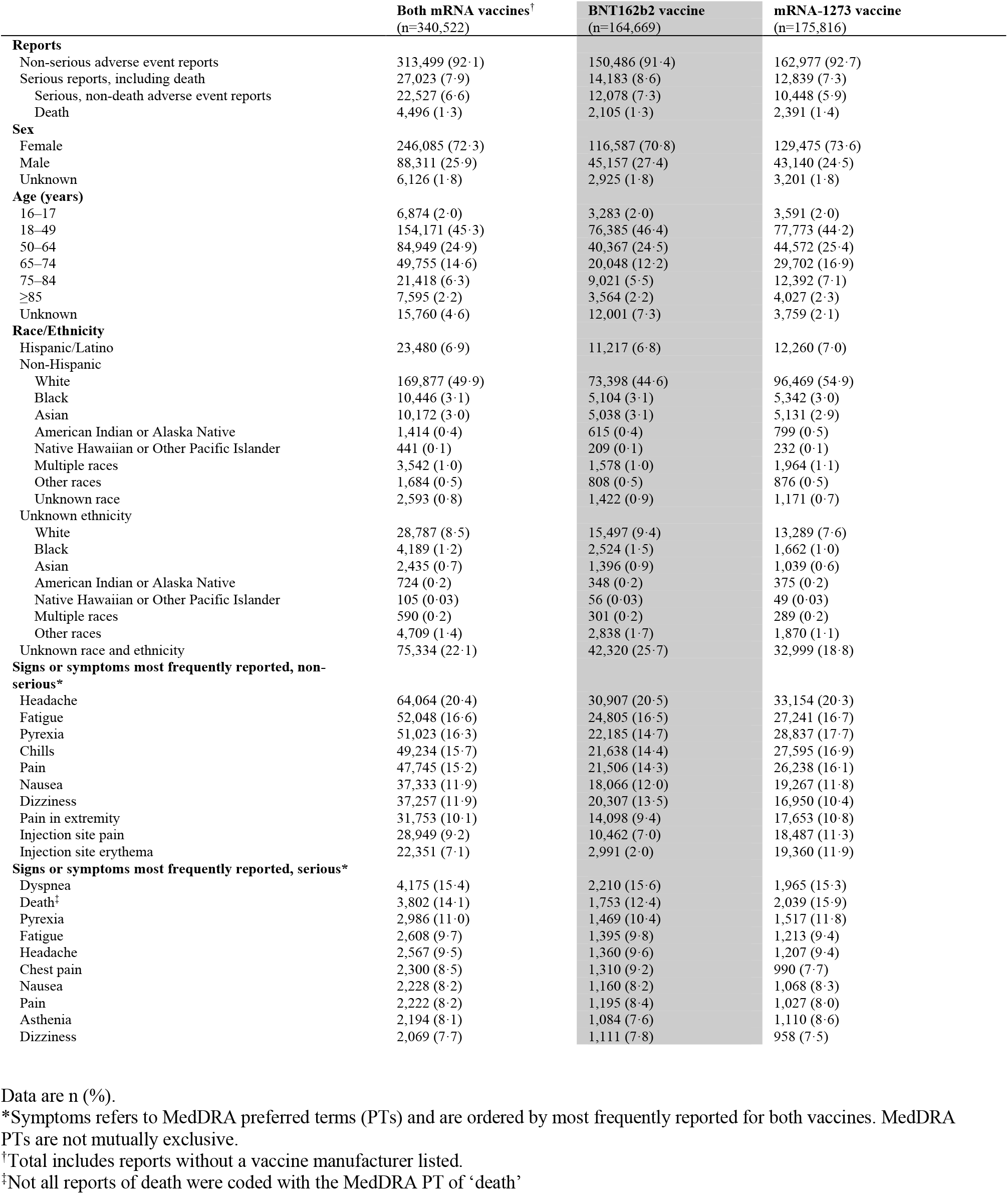
Characteristics of reports received and processed by Vaccine Adverse Events Reporting System (VAERS) for mRNA COVID-19 vaccines—December 14, 2020–June 14, 2021.

|  | Both mRNA vaccines<sup>†</sup><br>(n=340,522) | BNT162b2 vaccine<br>(n=164,669) | mRNA-1273 vaccine<br>(n=175,816) |
| --- | --- | --- | --- |
| **Reports** |  |  |  |
| Non-serious adverse event reports | 313,499 (92.1) | 150,486 (91.4) | 162,977 (92.7) |
| Serious reports, including death | 27,023 (7.9) | 14,183 (8.6) | 12,839 (7.3) |
| Serious, non-death adverse event reports | 22,527 (6.6) | 12,078 (7.3) | 10,448 (5.9) |
| Death | 4,496 (1.3) | 2,105 (1.3) | 2,391 (1.4) |
| **Sex** |  |  |  |
| Female | 246,085 (72.3) | 116,587 (70.8) | 129,475 (73.6) |
| Male | 88,311 (25.9) | 45,157 (27.4) | 43,140 (24.5) |
| Unknown | 6,126 (1.8) | 2,925 (1.8) | 3,201 (1.8) |
| **Age (years)** |  |  |  |
| 16–17 | 6,874 (2.0) | 3,283 (2.0) | 3,591 (2.0) |
| 18–49 | 154,171 (45.3) | 76,385 (46.4) | 77,773 (44.2) |
| 50–64 | 84,949 (24.9) | 40,367 (24.5) | 44,572 (25.4) |
| 65–74 | 49,755 (14.6) | 20,048 (12.2) | 29,702 (16.9) |
| 75–84 | 21,418 (6.3) | 9,021 (5.5) | 12,392 (7.1) |
| ≥85 | 7,595 (2.2) | 3,564 (2.2) | 4,027 (2.3) |
| Unknown | 15,760 (4.6) | 12,001 (7.3) | 3,759 (2.1) |
| **Race/Ethnicity** |  |  |  |
| Hispanic/Latino | 23,480 (6.9) | 11,217 (6.8) | 12,260 (7.0) |
| Non-Hispanic |  |  |  |
| White | 169,877 (49.9) | 73,398 (44.6) | 96,469 (54.9) |
| Black | 10,446 (3.1) | 5,104 (3.1) | 5,342 (3.0) |
| Asian | 10,172 (3.0) | 5,038 (3.1) | 5,131 (2.9) |
| American Indian or Alaska Native | 1,414 (0.4) | 615 (0.4) | 799 (0.5) |
| Native Hawaiian or Other Pacific Islander | 441 (0.1) | 209 (0.1) | 232 (0.1) |
| Multiple races | 3,542 (1.0) | 1,578 (1.0) | 1,964 (1.1) |
| Other races | 1,684 (0.5) | 808 (0.5) | 876 (0.5) |
| Unknown race | 2,593 (0.8) | 1,422 (0.9) | 1,171 (0.7) |
| Unknown ethnicity |  |  |  |
| White | 28,787 (8.5) | 15,497 (9.4) | 13,289 (7.6) |
| Black | 4,189 (1.2) | 2,524 (1.5) | 1,662 (1.0) |
| Asian | 2,435 (0.7) | 1,396 (0.9) | 1,039 (0.6) |
| American Indian or Alaska Native | 724 (0.2) | 348 (0.2) | 375 (0.2) |
| Native Hawaiian or Other Pacific Islander | 105 (0.03) | 56 (0.03) | 49 (0.03) |
| Multiple races | 590 (0.2) | 301 (0.2) | 289 (0.2) |
| Other races | 4,709 (1.4) | 2,838 (1.7) | 1,870 (1.1) |
| Unknown race and ethnicity | 75,334 (22.1) | 42,320 (25.7) | 32,999 (18.8) |
| **Signs or symptoms most frequently reported, non-serious\*** |  |  |  |
| Headache | 64,064 (20.4) | 30,907 (20.5) | 33,154 (20.3) |
| Fatigue | 52,048 (16.6) | 24,805 (16.5) | 27,241 (16.7) |
| Pyrexia | 51,023 (16.3) | 22,185 (14.7) | 28,837 (17.7) |
| Chills | 49,234 (15.7) | 21,638 (14.4) | 27,595 (16.9) |
| Pain | 47,745 (15.2) | 21,506 (14.3) | 26,238 (16.1) |
| Nausea | 37,333 (11.9) | 18,066 (12.0) | 19,267 (11.8) |
| Dizziness | 37,257 (11.9) | 20,307 (13.5) | 16,950 (10.4) |
| Pain in extremity | 31,753 (10.1) | 14,098 (9.4) | 17,653 (10.8) |
| Injection site pain | 28,949 (9.2) | 10,462 (7.0) | 18,487 (11.3) |
| Injection site erythema | 22,351 (7.1) | 2,991 (2.0) | 19,360 (11.9) |
| **Signs or symptoms most frequently reported, serious\*** |  |  |  |
| Dyspnea | 4,175 (15.4) | 2,210 (15.6) | 1,965 (15.3) |
| Death<sup>‡</sup> | 3,802 (14.1) | 1,753 (12.4) | 2,039 (15.9) |
| Pyrexia | 2,986 (11.0) | 1,469 (10.4) | 1,517 (11.8) |
| Fatigue | 2,608 (9.7) | 1,395 (9.8) | 1,213 (9.4) |
| Headache | 2,567 (9.5) | 1,360 (9.6) | 1,207 (9.4) |
| Chest pain | 2,300 (8.5) | 1,310 (9.2) | 990 (7.7) |
| Nausea | 2,228 (8.2) | 1,160 (8.2) | 1,068 (8.3) |
| Pain | 2,222 (8.2) | 1,195 (8.4) | 1,027 (8.0) |
| Asthenia | 2,194 (8.1) | 1,084 (7.6) | 1,110 (8.6) |
| Dizziness | 2,069 (7.7) | 1,111 (7.8) | 958 (7.5) |

Data are n (%).
\*Symptoms refers to MedDRA preferred terms (PTs) and are ordered by most frequently reported for both vaccines. MedDRA PTs are not mutually exclusive.
<sup>†</sup>Total includes reports without a vaccine manufacturer listed.
<sup>‡</sup>Not all reports of death were coded with the MedDRA PT of ‘death’

**Table 2:** Frequency and reporting rates of adverse events of special interest reported to Vaccine Adverse Event Reporting System (VAERS) by recipients of mRNA COVID-19 vaccine—December 14, 2020–June 14, 2021.

|  | Both mRNA vaccines |  | BNT162b2 vaccine |  | mRNA-1273 vaccine |  |
| --- | --- | --- | --- | --- | --- | --- |
|  | n | Reports per million doses administered* | n | Reports per million doses administered† | n | Reports per million doses administered‡ |
| Non-serious adverse event reports | 313,499 | 1,049.2 | 150,486 | 900.2 | 162,977 | 1,238.1 |
| Serious reports, including death | 27,023 | 90.4 | 14,183 | 84.8 | 12,839 | 97.5 |
| Serious, non-death adverse event reports | 22,527 | 75.4 | 12,078 | 72.2 | 10,448 | 79.4 |
| **Reports§ of adverse events of special interest\*\*** |  |  |  |  |  |  |
| COVID-19 | 9,344 | 31.3 | 7,184 | 43.0 | 2,160 | 16.4 |
| Coagulopathy†† | 4,320 | 14.5 | 2,343 | 14.0 | 1,977 | 15.0 |
| Seizure | 2,733 | 9.1 | 1,478 | 8.8 | 1,255 | 9.5 |
| Stroke‡‡ | 1,937 | 6.5 | 981 | 5.9 | 955 | 7.3 |
| Bells' Palsy | 1,918 | 6.4 | 1,057 | 6.3 | 861 | 6.5 |
| Anaphylaxis | 1,639 | 5.5 | 972 | 5.8 | 667 | 5.1 |
| Myopericarditis | 1,307 | 4.4 | 813 | 4.9 | 494 | 3.8 |
| Acute Myocardial Infarction | 1,118 | 3.7 | 610 | 3.6 | 508 | 3.9 |
| Appendicitis | 383 | 1.3 | 258 | 1.5 | 125 | 1.0 |
| Guillain-Barré Syndrome | 293 | 1.0 | 154 | 0.9 | 139 | 1.1 |
| Multisystem Inflammatory Syndrome in Adults | 119 | 0.4 | 60 | 0.4 | 59 | 0.4 |
| Transverse Myelitis | 98 | 0.3 | 55 | 0.3 | 43 | 0.3 |
| Narcolepsy | 21 | 0.1 | 12 | 0.1 | 9 | 0.1 |

\*298,792,852 doses of mRNA vaccine were administered in the study period.
†167,177,332 doses of BNT162b2 vaccine were administered in the study period.
‡131,639,515 doses of mRNA-1273 vaccine were administered in the study period.
§These represent reports, not confirmed by case definition.
\*\*Reported death is an adverse event of special interest but counts appear in following tables. Events are not mutually exclusive.
††Coagulopathy is an aggregate term capturing three specific adverse events: 1) thrombocytopenia, 2) deep venous thrombosis/pulmonary embolism, and 3) disseminated intravascular coagulopathy.
‡‡No vaccine manufacturer was provided for one report of stroke.

There were 4,496 reports of death to VAERS (Table 3). After review, 24 reports were excluded because of miscoding of death or duplicate reporting. Of the 4,472 reports of deaths analyzed, 2,087 (46·7%) were reported following BNT162b2 and 2,385 (53·3%) following mRNA-1273. Females accounted for 42·6% of reported deaths; the median age was 76 years (IQR: 66–86). More than 80% of deaths were reported among individuals aged 60 years or older (deaths reported per million doses administered by age group: 60–69 years, 14·4; 70–79 years, 28·5; 80–89 years, 75·4; ≥90 years, 2·1). 18·3% of decedents were identified as long-term care facility residents. Time interval to death following vaccination was available for 4,119 reports (92·1%); median time interval was 10·0 days (range: 0—161 days). The greatest number of death reports occurred on day 1 (10·5%) and day 2 (7·0%) following vaccination (Supplemental Figure 1). Compared to expected background rates of death from all causes per million vaccinated persons^20^, deaths reported to VAERS following mRNA vaccination were consistently 15-30 times less frequent within 7 days of vaccination, and 50 times less frequent within 42 days of vaccination, by age (Table 4).

**Table 3:**
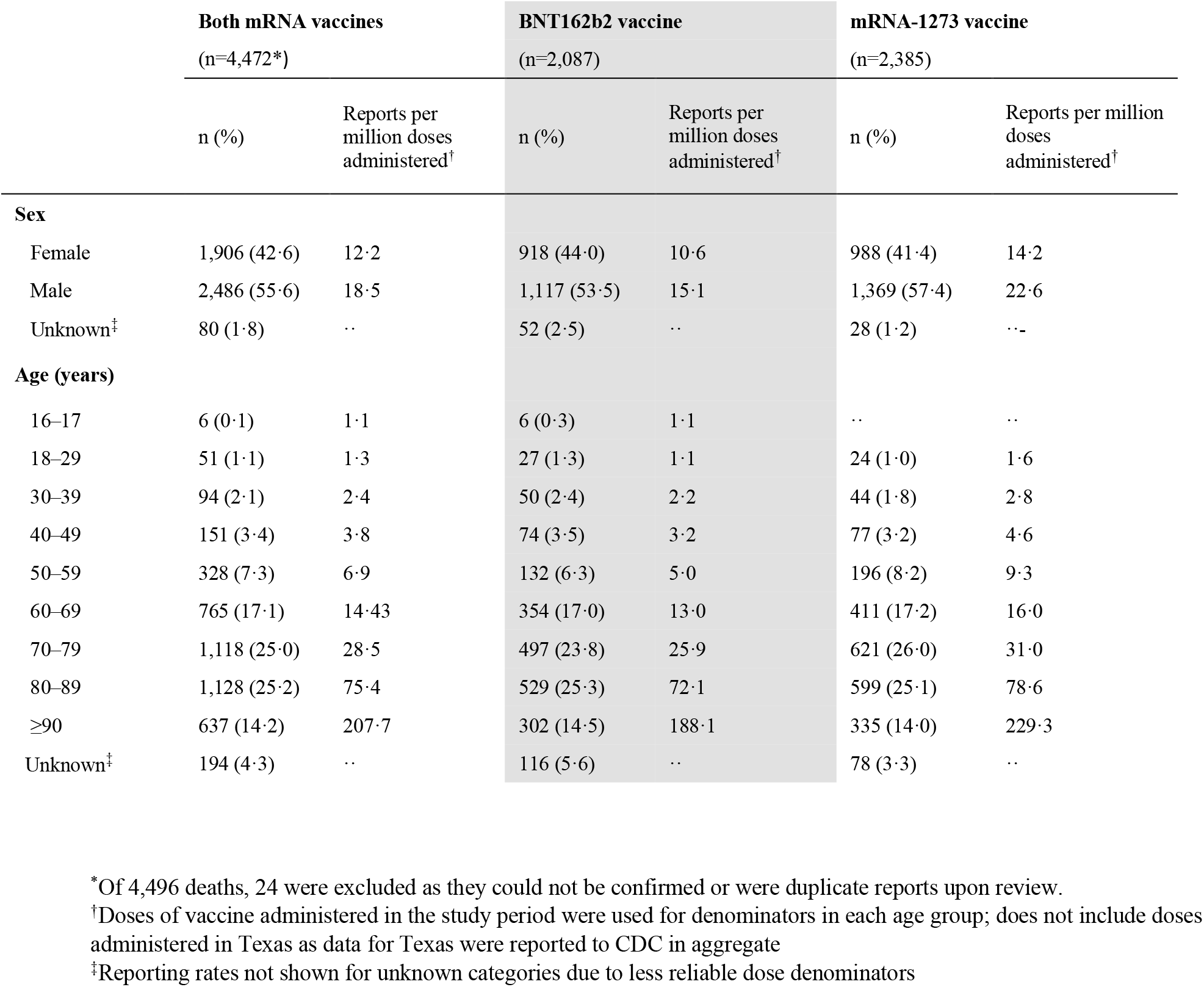
Frequency and reporting rates of death reported to Vaccine Adverse Event Reporting System (VAERS) by recipients of mRNA COVID-19 vaccine, by sex and age group—December 14, 2020–June 14, 2021.

**Table 4:** Expected rates of death per million vaccinated persons* and reported deaths to VAERS per million vaccinated persons within 7 and 42 days of vaccination among mRNA COVID-19 vaccine recipients, by age—December 14, 2020–June 14, 2021.

|  | Expected death rates |  | Rates based on reported deaths in VAERS |  |  |  |  |  |
| --- | --- | --- | --- | --- | --- | --- | --- | --- |
|  | All-cause deaths per million vaccinated persons |  | Both mRNA vaccines per million vaccinated persons |  | BNT162b2 vaccine per million vaccinated persons |  | mRNA-1273 vaccine per million vaccinated persons |  |
|  | Within 7 days of vaccination | Within 42 days of vaccination | Within 7 days of vaccination | Within 42 days of vaccination | Within 7 days of vaccination | Within 42 days of vaccination | Within 7 days of vaccination | Within 42 days of vaccination |
| **Age (years)<sup>†</sup>** |  |  |  |  |  |  |  |  |
| 16–24<sup>‡</sup> | 14.2 | 85.1 | 0.9 | 1.7 | 0.8 | 1.6 | 1.0 | 1.9 |
| 25–34 | 25.5 | 152.7 | 1.6 | 2.8 | 1.5 | 2.6 | 1.8 | 3.2 |
| 35–44 | 37.4 | 224.5 | 2.5 | 4.6 | 2.3 | 3.8 | 2.9 | 5.7 |
| 45–54 | 77.0 | 461.7 | 5.2 | 8.8 | 4.3 | 7.0 | 6.4 | 11.1 |
| 55–64 | 169.8 | 1,018.6 | 9.0 | 16.6 | 6.7 | 13.5 | 11.8 | 20.2 |
| 65–74 | 343.2 | 2,059.4 | 17.9 | 34.4 | 16.5 | 30.6 | 19.3 | 38.0 |
| 75–84 | 857.2 | 5,143.0 | 30.0 | 66.1 | 26.8 | 59.1 | 33.0 | 72.9 |
| ≥85 | 2,601.4 | 15,608.3 | 105.5 | 236.6 | 95.9 | 212.8 | 115.4 | 261.3 |
| **Total** | 165.6 | 993.3 | 11.7 | 23.6 | 9.2 | 18.5 | 15.0 | 30.5 |

\*Expected rates of deaths were derived from Abara WE, Gee J, Mu Y, Deloray M, Ye T, Shay DK, et al. *Expected Rates of Select Adverse Events following Immunization for COVID-19 Vaccine Safety Monitoring*. medRxiv. 2021 .
<sup>†</sup>Age grouping were selected for comparison to the expected rates age groups
<sup>‡</sup> The expected rates are among individuals aged 15-24

Death certificates or autopsy reports were available for clinical review for 808 (18·1%) reports of deaths analyzed. Among these, causes of death were most commonly diseases of the heart (46·5%) and COVID-19 disease (12·6%) (Supplemental Table 2, Supplemental Table 3). Among the 3,664 reports of death without a death certificate or autopsy, causes of death were most commonly unknown/unclear (54·1%), diseases of the heart (17·0%), and COVID-19 disease (8·7%). Causes of death by age among reports with death certificate or autopsy reports available are shown by age (Supplemental Table 4).

No adverse health outcome alerts were identified in EB data mining. However, five mRNA COVID-19 administrative error alerts (e.g. ‘product temperature excursion issue’) with disproportionality (EB05>2) were identified during the surveillance period.

### v-safe

During the analytic period, 7,914,583 mRNA COVID-19 vaccine recipients enrolled in v-safe and completed at least one post-vaccination health survey during days 0–7 (Table 5). The median age of v-safe participants was 50 years (IQR: 36–63), 62·9% were female, and 59·4% identified as non-Hispanic White. A total of 6,775,515 participants completed at least one survey during day 0–7 after dose 1 (3,455,778 following BNT162b2; 3,319,737 following mRNA-1273). Of these participants, 68·6% reported a local injection site reaction and 52·7% reported a systemic reaction. Of the 5,674,420 participants who completed surveys after dose 2, a greater percentage reported an injection site reaction (71·7%) and/or a systemic reaction (70·8%) (Table 6). Local injection site reactions were reported more frequently after mRNA-1273 (dose 1: 73·3%; dose 2: 78·4%) than after BNT162b2 (dose 1: 64·0%; dose 2: 65·3%). A similar pattern was found for systemic reactions after mRNA-1273 (dose 1: 54·3%; dose 2: 75·8%) versus BNT162b2 (dose 1: 51·3%; dose 2: 66·1%). The most frequently reported events after dose 1 of either mRNA vaccine included injection site pain (66·2%), fatigue (33·9%), and headache (27·0%); these were also more frequent after dose 2: injection site pain (68·6%), fatigue (55·7%), headache (46·2%). Differences in proportions of reactogenicity by dose number were similar after stratifying by age (<65 vs. ≥65 years) and sex. More reactogenicity was reported among younger participants aged <65 years and by females. (Supplemental Table 5).

**Table 5:** Demographic characteristics of v-safe participants reporting receipt of mRNA COVID-19 vaccine and completing at least one health survey 0-7 days after vaccination—December 14, 2020–June 14, 2021.

| Characteristics | Both mRNA vaccines | BNT162b2 vaccine |  | mRNA-1273 vaccine |  |
| --- | --- | --- | --- | --- | --- |
|  | (n = 7,914,583) | Dose 1<br>n=3,455,778 | Dose 2<br>n=2,920,526 | Dose 1<br>n= 3,319,737 | Dose 2<br>n=2,753,894 |
| **Sex** |  |  |  |  |  |
| Female | 4,975,209 (62.9) | 2,150,068 (62.2) | 1,861,599 (63.7) | 2,073,542 (62.5) | 1,779,200 (64.6) |
| Male | 2,860,738 (36.1) | 1,272,011 (36.8) | 1,032,941 (35.4) | 1,210,622 (36.5) | 947,612 (34.4) |
| Other | 8,872 (0.1) | 4,027 (0.1) | 3,464 (0.1) | 3,443 (0.1) | 2,947 (0.1) |
| Unknown | 69,764 (0.9) | 29,672 (0.9) | 22,522 (0.8) | 32,130 (1.0) | 24,135 (0.9) |
| **Age (years)** |  |  |  |  |  |
| 16–17 | 73,347 (0.9) | 63,865 (1.8) | 38,530 (1.3) | 946 (0.03) | 473 (0.02) |
| 18–49 | 3,791,839 (47.9) | 1,726,465 (50.0) | 1,431,627 (49.0) | 1,505,760 (45.4) | 1,219,210 (44.3) |
| 50–59 | 1,500,981 (19.0) | 653,799 (18.9) | 574,422 (19.7) | 627,214 (18.9) | 531,200 (19.3) |
| 60–64 | 739,381 (9.3) | 315,404 (9.1) | 279,350 (9.6) | 316,768 (9.5) | 270,831 (9.8) |
| 65–74 | 1,344,721 (17.0) | 516,227 (14.9) | 452,928 (15.5) | 643,663 (19.4) | 557,279 (20.2) |
| ≥75 | 464,314 (5.9) | 180,018 (5.2) | 143,669 (4.9) | 225,386 (6.8) | 174,901 (6.4) |
| **Race/Ethnicity** |  |  |  |  |  |
| Hispanic | 782,301 (9.9) | 346,197 (10.0) | 288,263 (9.9) | 316,460 (9.5) | 256,185 (9.3) |
| Non-Hispanic |  |  |  |  |  |
| White | 4,701,715 (59.4) | 2,059,560 (59.6) | 1,896,823 (64.9) | 1,979,056 (59.6) | 1,830,413 (66.5) |
| Black | 443,938 (5.6) | 202,598 (5.9) | 176,164 (6.0) | 178,981 (5.4) | 153,667 (5.6) |
| Asian | 467,932 (5.9) | 215,713 (6.2) | 196,173 (6.7) | 154,498 (4.7) | 138,793 (5.0) |
| American Indian or Alaska Native | 27,899 (0.4) | 11,161 (0.3) | 9,194 (0.3) | 13,486 (0.4) | 11,410 (0.4) |
| Native Hawaiian or Other Pacific Islander | 19,393 (0.2) | 8,500 (0.2) | 7,373 (0.3) | 7,689 (0.2) | 6,664 (0.2) |
| Multiple races | 110,326 (1.4) | 50,954 (1.5) | 46,129 (1.6) | 41,977 (1.3) | 38,772 (1.4) |
| Other races | 42,230 (0.5) | 19,252 (0.6) | 16,757 (0.6) | 15,885 (0.5) | 13,880 (0.5) |
| Unknown race | 23,420 (0.3) | 10,249 (0.3) | 9,090 (0.3) | 9,502 (0.3) | 8,270 (0.3) |
| Unknown ethnicity* |  |  |  |  |  |
| White | 115,766 (1.5) | 48,084 (1.4) | 38,674 (1.3) | 52,143 (1.6) | 42,070 (1.5) |
| Black | 26,865 (0.3) | 11,602 (0.3) | 8,570 (0.3) | 11,993 (0.4) | 8,406 (0.3) |
| Asian | 33,146 (0.4) | 14,134 (0.4) | 11,844 (0.4) | 11,356 (0.3) | 9,153 (0.3) |
| American Indian or Alaska Native | 3,142 (0.04) | 1,206 (0.03) | 848 (0.03) | 1,582 (0.05) | 1,151 (0.04) |
| Native Hawaiian or Other Pacific Islander | 1,945 (0.02) | 815 (0.02) | 659 (0.02) | 800 (0.02) | 613 (0.02) |
| Multiple races | 6,370 (0.1) | 2,902 (0.1) | 2,408 (0.1) | 2,478 (0.1) | 2,041 (0.1) |
| Other races | 13,148 (0.2) | 5,681 (0.2) | 4,528 (0.2) | 5,414 (0.2) | 4,263 (0.2) |
| Unknown race and ethnicity* | 129,647 (1.6) | 56,481 (1.6) | 45,410 (1.6) | 54,969 (1.7) | 44,340 (1.6) |
| Unavailable† | 965,400 (12.2) | 390,689 (11.3) | 161,619 (5.5) | 461,468 (13.9) | 183,803 (6.7) |
| **Pregnant at time of vaccination** | 86,801 (1.1) | 39,884 (1.2) | 39,163 (1.3) | 25,255 (0.8) | 25,428 (0.9) |
| **Pregnancy test positive after vaccination** | 27,370 (0.3) | 1,548 (0.04) | 11,677 (0.4) | 4,009 (0.1) | 10,199 (0.4) |

Data are n (%) unless otherwise stated.
\*Unknown indicates that v-safe participants selected unknown or preferred not to say.
†Unavailable refers to information that was not collected or missing in v-safe.

**Table 6:** Reported local and systemic reactions*, and reported health impact following mRNA COVID-19 vaccines reported days 0–7 after vaccination to v-safe, by manufacturer and dose—December 14, 2020 – June 14, 2021.

|  | Both mRNA vaccines |  | BNT162b2 vaccine |  | mRNA-1273 vaccine |  |
| --- | --- | --- | --- | --- | --- | --- |
|  | Dose 1<br>(n=6,775,515) | Dose 2<br>(n=5,674,420) | Dose 1<br>(n=3,455,778) | Dose 2<br>(n=2,920,526) | Dose 1<br>(n=3,319,737) | Dose 2<br>(n=2,753,894) |
| **Any injection site reaction** | 4,644,989 (68·6) | 4,068,447 (71·7) | 2,212,051 (64·0) | 1,908,124 (65·3) | 2,432,938 (73·3) | 2,160,323 (78·4) |
| Injection site pain | 4,488,402 (66·2) | 3,890,848 (68·6) | 2,140,843 (61·9) | 1,835,398 (62·8) | 2,347,559 (70·7) | 2,055,450 (74·6) |
| Swelling | 703,790 (10·4) | 976,946 (17·2) | 246,230 (7·1) | 309,718 (10·6) | 457,560 (13·8) | 667,228 (24·2) |
| Redness | 353,788 (5·2) | 640,739 (11·3) | 116,108 (3·4) | 167,127 (5·7) | 237,680 (7·2) | 473,612 (17·2) |
| Itching | 376,076 (5·6) | 605,633 (10·7) | 145,596 (4·2) | 191,132 (6·5) | 230,480 (6·9) | 414,501 (15·1) |
| **Any systemic reaction** | 3,573,429 (52·7) | 4,018,920 (70·8) | 1,771,509 (51·3) | 1,931,643 (66·1) | 1,801,920 (54·3) | 2,087,277 (75·8) |
| Fatigue | 2,295,205 (33·9) | 3,158,299 (55·7) | 1,127,904 (32·6) | 1,475,646 (50·5) | 1,167,301 (35·2) | 1,682,653 (61·1) |
| Headache | 1,831,471 (27·0) | 2,623,721 (46·2) | 893,992 (25·9) | 1,189,444 (40·7) | 937,479 (28·2) | 1,434,277 (52·1) |
| Myalgia | 1,423,336 (21·0) | 2,478,170 (43·7) | 653,821 (18·9) | 1,085,365 (37·2) | 769,515 (23·2) | 1,392,805 (50·6) |
| Chills | 631,546 (9·3) | 1,680,185 (29·6) | 263,617 (7·6) | 642,856 (22·0) | 367,929 (11·1) | 1,037,329 (37·7) |
| Fever | 642,092 (9·5) | 1,679,577 (29·6) | 274,650 (7·9) | 656,454 (22·5) | 367,442 (11·1) | 1,023,123 (37·2) |
| Joint pain | 642,006 (9·5) | 1,440,927 (25·4) | 285,812 (8·3) | 591,877 (20·3) | 356,194 (10·7) | 849,050 (30·8) |
| Nausea | 562,273 (8·3) | 901,103 (15·9) | 267,160 (7·7) | 384,525 (13·2) | 295,113 (8·9) | 516,578 (18·8) |
| Diarrhea | 383,576 (5·7) | 419,044 (7·4) | 190,542 (5·5) | 198,618 (6·8) | 193,034 (5·8) | 220,426 (8·0) |
| Abdominal pain | 233,511 (3·4) | 359,107 (6·3) | 113,872 (3·3) | 158,251 (5·4) | 119,639 (3·6) | 200,856 (7·3) |
| Rash | 85,766 (1·3) | 99,878 (1·8) | 41,565 (1·2) | 42,662 (1·5) | 44,201 (1·3) | 57,216 (2·1) |
| Vomiting | 55,710 (0·8) | 91,727 (1·6) | 25,336 (0·7) | 36,761 (1·3) | 30,374 (0·9) | 54,966 (2·0) |
| **With reported health impact\*** | 808,963 (11·9) | 1,821,421 (32·1) | 361,834 (10·5) | 740,529 (25·4) | 447,129 (13·5) | 1,080,892 (39·2) |
| Unable to do normal activity | 658,330 (9·7) | 1,501,679 (26·5) | 290,207 (8·4) | 598,584 (20·5) | 368,123 (11·1) | 903,095 (32·8) |
| Unable to work | 305,709 (4·5) | 911,366 (16·1) | 135,063 (3·9) | 360,411 (12·3) | 170,646 (5·1) | 550,955 (20·0) |
| Reported medical care | 56,647 (0·8) | 53,077 (0·9) | 27,358 (0·8) | 25,568 (0·9) | 29,289 (0·9) | 27,509 (1·0) |
| Telehealth | 19,562 (0·3) | 19,770 (0·3) | 9,318 (0·3) | 9,238 (0·3) | 10,244 (0·3) | 10,532 (0·4) |
| Clinic | 18,671 (0·3) | 16,793 (0·3) | 9,109 (0·3) | 8,487 (0·3) | 9,562 (0·3) | 8,306 (0·3) |
| Emergency visit | 9,907 (0·1) | 8,907 (0·2) | 5,087 (0·1) | 4,494 (0·2) | 4,820 (0·1) | 4,413 (0·2) |
| Hospitalization | 1,896 (0·03) | 2,053 (0·04) | 915 (0·03) | 1,001 (0·03) | 981 (0·03) | 1,052 (0·04) |

Data are n (%) unless otherwise stated.
\*Reports of local and systemic reactions, and reports of health impact are not mutually exclusive.

Local and systemic reactions by manufacturer, dose, days after vaccination, and severity are shown in Figure 1. The majority of reported symptoms were mild. Participants reported moderate and severe reactogenicity most commonly on day 1 after dose 2 of either vaccine. The proportion of participants who reported symptoms was greatest on day 1 and then decreased subsequently. The highest proportion of participants reported severe symptoms on day 1 following dose 2 of mRNA-1273 (Supplemental Table 7). On all other days, proportions of participants reporting severe symptoms did not exceed 3.0% for any individual symptom (Supplemental Tables 6 and 7).

**Figure 1:**
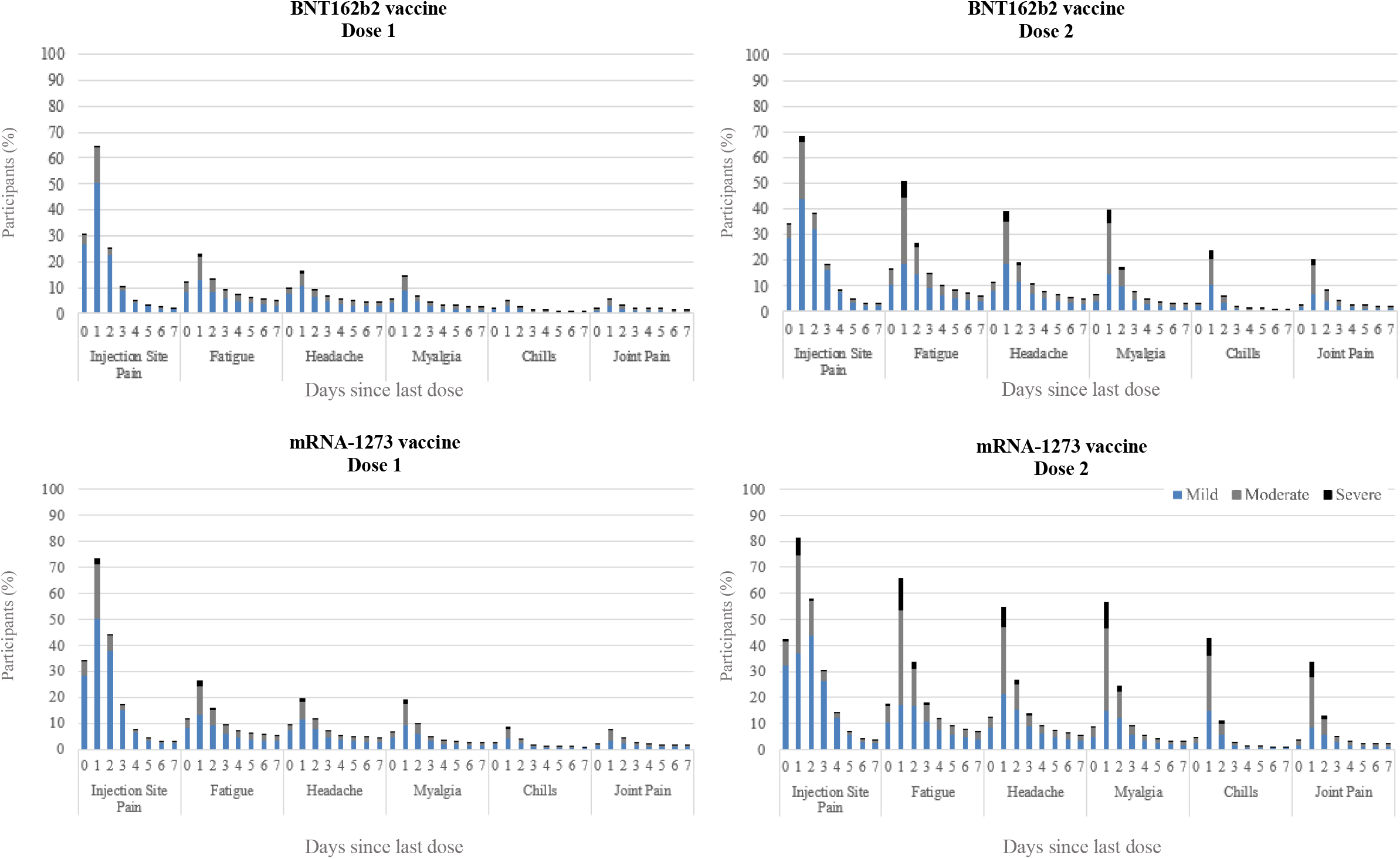
Local and systemic reactions^*^ to mRNA COVID-19 vaccine reported in v-safe, by manufacturer, dose, days after vaccination, and severity^†^. *Top five reactions determined by reported frequency after dose 2 of both mRNA COVID-19 vaccines in v-safe, excluding fever because it was not rated mild, moderate, or severe. ^†^Mild was defined as “noticeable symptoms but they aren’t a problem”, moderate was defined as “symptoms that limit normal activities, and severe symptoms “make normal daily activities difficult or impossible”

Reported health impact was greater following dose 2 of either vaccine (32·1%) compared with dose 1 (11·9%) and after mRNA-1273 of either dose compared with BNT162b2 (Table 6). After dose 2 of BNT162b2, 20·5% were unable to do normal activities, and 12·3% were unable to work. After dose 2 of mRNA-1273, 32·8% were unable to do normal activities, and 20·0% were unable to work. Less than 1·0% reported receiving medical care after receiving either dose of either vaccine. Fewer participants reported an emergency room visit (dose 1: 0·1%; dose 2: 0·2%) or hospitalization (dose 1: 0·03%; dose 2: 0·04%).

When stratified by sex, females reported a health impact more frequently than males, peaking on day 1 after vaccination (Supplemental Figure 2). Following dose 2 of mRNA-1273 vaccine, 41·4% of females reported an inability to perform normal activities in the day 1 survey, and 23·5% an inability to work. Among males receiving dose 2 of mRNA-1273 on the day 1 survey, 25·6% were unable to perform normal activity and 16·9% were unable to work (Supplemental Table 8).

## Discussion

In this analysis of the first six months of VAERS and v-safe data, when over 298 million doses of mRNA vaccines were administered, we report reactogenicity following mRNA COVID-19 vaccination, and reported adverse events, including deaths. Reactogenicity was similar to what was reported from clinical trials and from early post-authorization monitoring.^3-5,10,11^ In both VAERS and v-safe, local injection site and systemic reactions were commonly reported. V-safe participants more frequently reported transient reactions following mRNA-1273 compared with BNT162b2, and more frequently following dose 2 of either vaccine. Females and individuals aged <65 years reported AEs and reactions more frequently. Analysis of deaths reported to VAERS demonstrated lower than expected reported mortality rates compared with background mortality rates.

Safety monitoring of COVID-19 vaccines has been the most comprehensive in recent U.S. history. COVID-19 vaccine safety has been monitored with established systems, including the Vaccine Safety Datalink (VSD)^23^ and VAERS, and a new system, v-safe, developed specifically for COVID-19 vaccine safety monitoring. During the study period, all COVID-19 vaccines were administered under EUAs, which require vaccine providers to report serious safety events that follow vaccination to VAERS. Heightened public awareness of the COVID-19 vaccination program, outreach and education to healthcare providers and hospitals about COVID-19 EUA AE reporting requirements, and adherence to EUA reporting requirements by providers and health systems, are likely contributing to the high volume of VAERS reports following mRNA vaccination. Data from U.S. safety monitoring systems have been reviewed regularly by the ACIP COVID-19 Vaccine Safety Technical (VaST) work group^24^ and at public ACIP meetings.^25^ Specific safety evaluations have covered serious AEs including anaphylaxis,^14^ thrombosis with thrombocytopenia syndrome (TTS),^26^ myocarditis,^27^ and Guillain-Barré Syndrome (GBS).^28^ ACIP has assessed the benefit-risk balance of each of the currently licensed or authorized U.S. COVID-19 vaccines; these evaluations have not prompted any changes in U.S. COVID-19 vaccine recommendations.^27,28^

Reactogenicity findings from VAERS and v-safe were similar to those from a large study of reactogenicity conducted in the United Kingdom.^29^ The observed patterns may be explained in part by host characteristics known to influence reactogenicity, including age, sex, and the presence of underlying medical conditions.^30^ Females have more vigorous antibody responses^31^ to certain vaccines and also tend to report more severe local and systemic reactions to influenza vaccine.^32^ Females may also be more likely than males to respond to surveys.^33,34^ Younger persons may be more comfortable with smartphone-based surveys and more likely to respond to surveys, generally.^35,36^

Using v-safe data, we were able to assess the effects of mRNA vaccination on daily life activities for the first time in adults. Effects on daily activities were most frequently reported on day 1 after vaccination. Reports about the health impact measures used in v-safe, while self-assessed and subjective, correlate with reports about reactogenicity: more health impact was reported by females than males, by participants aged <65 years compared with older participants, after dose 2 compared with dose 1, and by those who received mRNA-1273 versus BNT162b2. Reports of seeking medical care after mRNA vaccine were rare; v-safe did not ask which symptoms prompted the participant to seek medical care. Reactogenicity and its associated health effects, even if transient, may deter some from seeking vaccination. Surveys found that nearly half of unvaccinated adults aged <50 years expressed concern about missing work due to vaccine side effects and that employees provided time off were more likely to get vaccinated;^37,38^ employer policies that accommodate this might increase vaccination coverage.^39^

Patterns of VAERS reports following mRNA COVID-19 vaccination were similar to reports following receipt of other vaccines routinely administered to adults; most reported events were non-serious.^40-43^ EB data mining findings did not suggest any unexpected vaccine safety problems based on pre-specified statistical thresholds. Serious AEs have been detected following receipt of COVID-19 vaccines during U.S. safety monitoring and reviewed in detail in other reports.^26-28^ Reports of anaphylaxis and myocarditis following mRNA vaccination prompted clinical guidance and management recommendations.^27,44-47^ TTS^26^ and GBS^28^ have been associated with Janssen COVID-19 vaccination (Ad26.COV2.S) but not with mRNA vaccination to date.

In our review and analysis of death reports to VAERS following mRNA vaccination, we found no unusual patterns in cause of death among the large number of death reports received. Reporting rates of death to VAERS among vaccinated persons in 7- and 42-day periods following vaccination were lower than expected background mortality rates. Under the COVID-19 vaccine EUA regulations, healthcare providers are required to report deaths and life-threatening potential adverse events that follow COVID-19 vaccinations to VAERS regardless of their potential association with vaccination. These requirements make comparing the number of reported deaths to VAERS for COVID-19 vaccines to reported deaths following other adult vaccines^48^ difficult. Older individuals and those in long-term care facilities with the highest baseline mortality risk were initially prioritized for COVID-19 vaccination while most U.S. immunization programs target pediatric and adolescent populations.^49^ The concentrated reporting of deaths on the first few days following vaccination follows patterns similar to those observed for other adult vaccinations.^48^ This pattern may represent reporting bias, as the likelihood to report a serious AE may increase when it occurs in close temporal proximity to vaccination.

There are clear limitations in any review of preliminary mortality data. A comparison to national mortality data,^20,50^ suggests that certain causes of death such as accidents, suicides, or cancer, are less likely to be reported to VAERS. The predominance of heart disease as a cause of death reports to VAERS warrants continued monitoring and assessment, but may be driven by non-specific causes, like cardiac arrest, that might be chosen as a terminal event if no immediate explanation is available. Death certificate or autopsy reports were available only for a small proportion of deaths reported to VAERS. Finally, VAERS is designed as an early warning system to detect potential safety signals^8^ and VAERS data alone generally cannot establish causal relationships between vaccination and AEs. Other studies and surveillance systems, including the VSD, are needed to better understand patterns of mortality following mRNA COVID-19 vaccination.^23,51^

This study has several strengths, including the large population under surveillance and comprehensive capture of national data from two complementary surveillance systems. Because the U.S. government purchased all COVID-19 doses and collected administration data, we were able to calculate VAERS reporting rates using the number of mRNA vaccine doses administered.^52^ In contrast, past VAERS analyses for other vaccines could only use doses distributed, which are always greater than doses administered as denominators, underestimating vaccine adverse event reporting rates. Data concerning how reactogenicity following mRNA vaccination affects daily activities and work during the week following vaccination from v-safe provide information has not been previously described. Limitations of this analysis include a limitation shared by all analyses using VAERS data: it is a passive reporting system subject to underreporting and variable or incomplete reporting.^8^ While VAERS death reports were individually reviewed by CDC physicians and follow-up is ongoing to obtain additional and missing records, other serious AE reports were not individually reviewed. Additionally, VAERS reports require interpretation to determine if AE reports meet clinical case definitions.^53^ EB data mining has multiple limitations,^21,22^ including that the absence of a disproportionality alert for an event does not rule out a possible corresponding adverse event. A new concern with disproportionality scores, which are adjusted by year to control for time-dependent confounders, is that during the study period most VAERS reports were for COVID-19 vaccinations. As all VAERS reports are used for vaccine-event comparisons in EB data mining, potential associations with mRNA COVID-19 vaccines plausibly could be missed. A limitation of v-safe is the need for smartphone access. Because a subset of all vaccine recipients chose or were able to participate in v-safe, the results likely are not generalizable to the entire vaccinated U.S population. In addition, participants in v-safe may be lost to follow up as there is no requirement for continuous enrollment. Finally, this report only included v-safe responses received during the first week post-vaccination.

During the first six months of the U.S. COVID-19 vaccination program, more than 50% of the eligible population received at least one vaccine dose.^19^ VAERS and v-safe data from this period demonstrate a post-authorization safety profile for mRNA COVID-19 vaccines that is generally consistent with pre-authorization trials^3,4^ and early post-authorization surveillance reports.^10,11^ Serious AEs have been identified following mRNA vaccinations; however, based on the most current information, these events are rare. Vaccines are the most effective tool to prevent serious COVID-19 disease outcomes^54^ and the benefits of immunization in preventing serious morbidity and mortality clearly favor vaccination.^26-28^ VAERS and v-safe, two complementary surveillance systems, will continue to provide data needed to inform immunization policy makers, medical and immunization providers, and the public about the safety of COVID-19 vaccination.

## Data Availability

Data produced in the present study are available upon reasonable request to the authors

## Acknowledgements

We wish to acknowledge the following contributors: CDC: Amelia Jazwa, Tara Johnson, Charles Licata, Stacey Martin, Florence Lee, Carla Black; FDA: Jane Baumblatt, Deborah Thompson, Kerry Welsh, Narayan Nair, Kosal Nguon (Commonwealth Informatics); v-safe participants; Oracle v-safe development team.

---

**Supplemental Table 1:** mRNA COVID-19 vaccine doses administered in the United States—December 14, 2020–June 14, 2021.

| Characteristics | Both mRNA vaccines<sup>†</sup><br>(n=298,792,852) | BNT162b2 vaccine<sup>†</sup><br>(n=167,177,332) | mRNA-1273 vaccine<sup>†</sup><br>(n=131,639,515) |
| --- | --- | --- | --- |
| **Sex** |  |  |  |
| Female | 155,969,573 (53.2) | 86,507,992 (53.5) | 69,461,582 (52.8) |
| Male | 134,373,958 (45.8) | 73,768,602 (45.6) | 60,605,356 (46.1) |
| Unknown | 2,868,979 (1.0) | 1,452,344 (0.9) | 1,416,634 (1.1) |
| **Age (years)** |  |  |  |
| 16–17* | 5,506,763 (1.8) | 5,365,855 (3.2) | 140,908 (0.1) |
| 18–49 | 126,288,626 (42.3) | 74,999,327 (44.9) | 51,289,299 (39.0) |
| 50–64 | 79,207,752 (26.5) | 43,595,972 (26.1) | 35,611,780 (27.1) |
| 65–74 | 51,699,307 (17.3) | 25,402,217 (15.2) | 26,297,090 (20.0) |
| 75–84 | 27,731,181 (9.3) | 13,555,128 (8.1) | 14,176,053 (10.8) |
| ≥85 | 8,359,223 (2.8) | 4,248,648 (2.5) | 4,110,575 (3.1) |
| Unknown | 23,995 (0.01) | 10,185 (0.01) | 13,810 (0.01) |
| **Race/Ethnicity** |  |  |  |
| Hispanic | 31,599,632 (10.8) | 17,964,345 (11.1) | 13,635,287 (10.4) |
| Non-Hispanic |  |  |  |
| White | 112,698,875 (38.4) | 61,996,607 (38.3) | 50,702,268 (38.6) |
| Asian | 11,789,429 (4.0) | 7,258,033 (4.5) | 4,531,396 (3.4) |
| Black | 16,848,436 (5.7) | 9,665,586 (6.0) | 7,182,849 (5.5) |
| American Indian or Alaska Native | 1,738,938 (0.6) | 842,263 (0.5) | 896,674 (0.7) |
| Native Hawaiian or Other Pacific Islander | 508,285 (0.2) | 295,634 (0.2) | 212,651 (0.2) |
| Multiple races | 8,856,800 (3.0) | 5,037,828 (3.1) | 3,818,972 (2.9) |
| Other races | 6,949,404 (2.4) | 4,161,353 (2.6) | 2,788,051 (2.1) |
| Unknown race and ethnicity | 102,227,532 (34.9) | 54,511,493 (33.7) | 47,716,039 (36.3) |

Data are n (%) unless otherwise stated.
\*mRNA-1273 vaccine was not authorized for individuals <18 years during this period, reported mRNA-1273 doses are either from clinical trials or were administered or reported in error
<sup>†</sup>Totals reflect the number of doses in age categories. Missing doses for sex and race/ethnicity are due to certain jurisdictions that report data in aggregate.

**Supplemental Table 2:** Most common causes of death among reports received and processed by Vaccine Adverse Event Reporting System (VAERS) following mRNA COVID-19 vaccination (n=4,472)—December 14, 2020–June 14, 2021.

| ICD-10 Major Group | Death or autopsy certificate available |  |  | No death certificate or autopsy available |  |  |
| --- | --- | --- | --- | --- | --- | --- |
|  | Both mRNA vaccines<br>(n=808) | BNT162b2 vaccine<br>(n=401) | mRNA-1273 vaccine<br>(n=407) | Both mRNA vaccines<br>(n=3,664) | BNT162b2 vaccine<br>(n=1,686) | mRNA-1273 vaccine<br>(n=1,978) |
| **All reported deaths** |  |  |  |  |  |  |
| Diseases of the heart | 376 (46.5) | 161 (40.1) | 215 (52.8) | 622 (17.0) | 296 (17.6) | 326 (16.5) |
| COVID-19 disease | 102 (12.6) | 62 (15.5) | 40 (9.8) | 317 (8.7) | 178 (10.6) | 139 (7.0) |
| Other | 68 (8.4) | 38 (9.5) | 30 (7.4) | 141 (3.8) | 68 (4.0) | 73 (3.7) |
| Cerebrovascular diseases | 53 (6.6) | 28 (7.0) | 25 (6.1) | 207 (5.6) | 101 (6.0) | 106 (5.4) |
| Dementia | 41 (5.1) | 20 (5.0) | 21 (5.2) | 9 (0.2) | 3 (0.2) | 6 (0.3) |
| Chronic lower respiratory diseases | 28 (3.5) | 17 (4.2) | 11 (2.7) | 29 (0.8) | 10 (0.6) | 19 (1.0) |
| Malignant neoplasms | 27 (3.3) | 15 (3.7) | 12 (2.9) | 68 (1.9) | 42 (2.5) | 26 (1.3) |
| Unknown/unclear | 27 (3.3) | 9 (2.2) | 18 (4.4) | 1,984 (54.1) | 844 (50.1) | 1,140 (57.6) |
| Septicemia | 23 (2.8) | 12 (3.0) | 11 (2.7) | 72 (2.0) | 47 (2.8) | 25 (1.3) |
| Influenza and pneumonia | 22 (2.7) | 18 (4.5) | 4 (1.0) | 113 (3.1) | 52 (3.1) | 61 (3.1) |
| Accidents/unintentional injuries | 11 (1.4) | 3 (0.7) | 8 (2.0) | 22 (0.6) | 8 (0.5) | 14 (0.7) |
| Renal disease | 8 (1.0) | 5 (1.2) | 3 (0.7) | 25 (0.7) | 7 (0.4) | 18 (0.9) |
| Hematologic disease, other than malignancy | 7 (0.9) | 5 (1.2) | 2 (0.5) | 19 (0.5) | 9 (0.5) | 10 (0.5) |
| Pneumonitis due to solids and liquids | 6 (0.7) | 3 (0.7) | 3 (0.7) | 8 (0.2) | 5 (0.3) | 3 (0.2) |
| Diabetes mellitus | 4 (0.5) | 1 (0.2) | 3 (0.7) | 6 (0.2) | 4 (0.2) | 2 (0.1) |
| Chronic liver disease and cirrhosis | 4 (0.5) | 3 (0.7) | 1 (0.2) | 7 (0.2) | 4 (0.2) | 3 (0.2) |
| Intentional self-harm | 1 (0.1) | 1 (0.2) | 0 (0.0) | 15 (0.4) | 8 (0.5) | 7 (0.4) |

Data are n (%) unless otherwise stated.

**Supplemental Table 3:** Causes and impressions of death among reported deaths to Vaccine Adverse Event Reporting System (VAERS) December 14, 2020–June 14, 2021 following mRNA vaccination.

| ICD-10 Major Group and impression | All reports of death | Reported deaths with death certificate or autopsy |
| --- | --- | --- |
| **All reported deaths, n** | 4,472 | 808 |
| Diseases of the heart | 998 (22.3) | 376 (46.5) |
| *Aortic dissection, aneurysm or aortitis* | 13 (0.3) | 7 (0.9) |
| *Arrhythmia* | 42 (0.9) | 18 (2.2) |
| *Atherosclerotic cardiovascular or hypertensive cardiovascular disease* | 129 (2.9) | 98 (12.1) |
| *Cardiac arrest* | 321 (7.2) | 79 (9.8) |
| *Cardiomyopathy or hypertrophy* | 17 (0.4) | 12 (1.5) |
| *Heart failure* | 104 (2.3) | 47 (5.8) |
| *Myocardial infarction* | 247 (5.5) | 87 (10.8) |
| *Myocarditis* | 4 (0.1) | 0 (0.0) |
| *Pulmonary embolism* | 84 (1.9) | 17 (2.1) |
| *Other cardiac cause* | 37 (0.8) | 11 (1.4) |
| COVID-19 disease | 419 (9.4) | 102 (12.6) |
| Cerebrovascular diseases | 260 (5.8) | 53 (6.6) |
| Other | 209 (4.7) | 68 (8.4) |
| *Disseminated herpes zoster* | 2 (0.04) | 1 (0.1) |
| *Drug overdose/intoxication* | 7 (0.2) | 5 (0.6) |
| *Failure to thrive* | 9 (0.2) | 9 (1.1) |
| *Gastrointestinal\** | 36 (0.8) | 8 (1.0) |
| *Hemorrhage/Hemorrhagic shock* | 4 (0.1) | 2 (0.2) |
| *Metabolic derangement* | 4 (0.1) | 1 (0.1) |
| *Multiorgan failure* | 28 (0.6) | 7 (0.9) |
| *Natural* | 2 (0.04) | 2 (0.2) |
| *Neurologic†* | 28 (0.6) | 6 (0.7) |
| *Obesity* | 2 (0.04) | 2 (0.2) |
| *Respiratory failure* | 63 (1.4) | 22 (2.7) |
| *Vaccine related‡* | 4 (0.1) | 3 (0.4) |
| Influenza and pneumonia | 135 (3.0) | 22 (2.7) |
| Malignant neoplasms | 95 (2.1) | 27 (3.3) |
| Septicemia | 95 (2.1) | 23 (2.8) |
| Chronic lower respiratory diseases | 57 (1.3) | 28 (3.5) |
| Dementia | 50 (1.1) | 41 (5.1) |
| Accidents/unintentional injuries | 33 (0.7) | 11 (1.4) |
| Renal disease | 33 (0.7) | 8 (1.0) |
| Hematologic disease, other than malignancy | 26 (0.6) | 7 (0.9) |
| Intentional self-harm | 16 (0.4) | 1 (0.1) |
| Pneumonitis due to solids and liquids | 14 (0.3) | 6 (0.7) |
| Chronic liver disease and cirrhosis | 11 (0.2) | 4 (0.5) |
| Diabetes mellitus | 10 (0.2) | 4 (0.5) |
| Unknown/unclear | 2,011 (45.0) | 27 (3.3) |

Data are n (%) unless otherwise stated.
\*Gastrointestinal includes gastrointestinal bleeding, bowel obstruction/perforation, mesenteric ischemia, pancreatitis.
†Neurologic includes amyotrophic lateral sclerosis, encephalopathy, hydrocephalus, Guillain-Barré syndrome, seizure.
‡Vaccine related includes systemic inflammatory response syndrome from vaccine reaction, anaphylaxis post-COVID-19 vaccination

**Supplemental Table 4:** Causes of death among reports of death with death certificate or autopsy available to Vaccine Adverse Event Reporting System (VAERS) December 14, 2020–June 14, 2021 following mRNA vaccination, by age.

| ICD-10 Major Group | All<br>808 | 16-24<br>n=4 |  | 25-34<br>n=5 |  | 35-44<br>n=24 |  | 45-54<br>n=37 |  | 55-64<br>n=101 |  | 65-74<br>n=171 |  | 75-84<br>n=202 |  | 85+<br>n=264 |  |
| --- | --- | --- | --- | --- | --- | --- | --- | --- | --- | --- | --- | --- | --- | --- | --- | --- | --- |
|  |  | n | % | n | % | n | % | n | % | n | % | n | % | n | % | n | % |
| Diseases of the heart | 376 | 1 | 25.0 | 2 | 40.0 | 10 | 41.7 | 21 | 56.8 | 50 | 49.5 | 98 | 57.3 | 91 | 45.0 | 103 | 39.0 |
| COVID-19 disease | 102 | 1 | 25.0 | 1 | 20.0 | 2 | 8.3 | 2 | 5.4 | 11 | 10.9 | 19 | 11.1 | 32 | 15.8 | 34 | 12.9 |
| Cerebrovascular diseases | 53 | 0 | 0 | 0 | 0 | 0 | 0 | 4 | 10.8 | 8 | 7.9 | 8 | 4.7 | 13 | 6.4 | 20 | 7.6 |
| Other | 68 | 0 | 0 | 1 | 20.0 | 5 | 20.8 | 2 | 5.4 | 10 | 9.9 | 11 | 6.4 | 15 | 7.4 | 24 | 9.1 |
| Influenza and pneumonia | 22 | 0 | 0 | 0 | 0 | 2 | 8.3 | 0 | 0 | 2 | 2.0 | 4 | 2.3 | 7 | 3.5 | 7 | 2.7 |
| Malignant neoplasms | 27 | 0 | 0 | 0 | 0 | 0 | 0 | 2 | 5.4 | 5 | 5.0 | 2 | 1.2 | 8 | 4.0 | 10 | 3.8 |
| Septicemia | 23 | 0 | 0 | 0 | 0 | 0 | 0 | 1 | 2.7 | 2 | 2.0 | 2 | 1.2 | 6 | 3.0 | 12 | 4.5 |
| Chronic lower respiratory diseases | 28 | 0 | 0 | 0 | 0 | 0 | 0 | 0 | 0 | 2 | 2.0 | 11 | 6.4 | 4 | 2.0 | 11 | 4.2 |
| Dementia, including Alzheimer's, Parkinson's disease | 41 | 0 | 0 | 0 | 0 | 0 | 0 | 0 | 0 | 1 | 1.0 | 1 | 0.6 | 7 | 3.5 | 32 | 12.1 |
| Accidents/unintentional injuries | 11 | 0 | 0 | 0 | 0 | 1 | 4.2 | 0 | 0 | 2 | 2.0 | 2 | 1.2 | 3 | 1.5 | 3 | 1.1 |
| Renal disease, including nephritis and chronic disease | 8 | 0 | 0 | 0 | 0 | 0 | 0 | 1 | 2.7 | 0 | 0 | 3 | 1.8 | 3 | 1.5 | 1 | 0.4 |
| Hematologic disease, other than malignancy | 7 | 0 | 0 | 0 | 0 | 0 | 0 | 1 | 2.7 | 3 | 3.0 | 0 | 0 | 1 | 0.5 | 2 | 0.8 |
| Intentional self-harm | 1 | 1 | 25.0 | 0 | 0 | 0 | 0 | 0 | 0 | 0 | 0 | 0 | 0 | 0 | 0 | 0 | 0 |
| Pneumonitis due to solids and liquids | 6 | 0 | 0 | 0 | 0 | 0 | 0 | 0 | 0 | 1 | 1.0 | 0 | 0 | 3 | 1.5 | 2 | 0.8 |
| Chronic liver disease and cirrhosis | 4 | 0 | 0 | 1 | 20.0 | 0 | 0 | 0 | 0 | 1 | 1.0 | 1 | 0.6 | 1 | 0.5 | 0 | 0 |
| Diabetes mellitus | 4 | 0 | 0 | 0 | 0 | 1 | 4.2 | 0 | 0 | 0 | 0 | 2 | 1.2 | 1 | 0.5 | 0 | 0 |
| Unknown/unclear | 27 | 1 | 25.0 | 0 | 0 | 3 | 12.5 | 3 | 8.1 | 3 | 3.0 | 7 | 4.1 | 7 | 3.5 | 3 | 1.1 |

**Supplemental Table 5:** Local and systemic reactions^*^ 0–7 days after vaccination by sex, age, and dose number, reported in v-safe—December 14, 2020– June 14, 2021.

|  | Female |  | Male |  | <65 years |  | ≥65 years |  |
| --- | --- | --- | --- | --- | --- | --- | --- | --- |
|  | Dose 1 | Dose 2 | Dose 1 | Dose 2 | Dose 1 | Dose 2 | Dose 1 | Dose 2 |
|  | (n=4,223,610) | (n=3,640,799) | (n=2,482,633) | (n=1,980,553) | (n=5,210,221) | (n=4,345,643) | (n=1,565,294) | (n=1,328,777) |
| **Any injection site reaction** | 3,095,194 (73·3) | 2,792,488 (76·7) | 1,498,108 (60·3) | 1,235,278 (62·4) | 3,835,618 (73·6) | 3,290,206 (75·7) | 809,371 (51·7) | 778,241 (58·6) |
| Injection site pain | 2,989,733 (70·8) | 2,666,734 (73·3) | 1,448,440 (58·3) | 1,184,914 (59·8) | 3,728,795 (71·6) | 3,179,024 (73·2) | 759,607 (48·5) | 711,824 (53·6) |
| Swelling | 539,793 (12·8) | 771,962 (21·2) | 154,980 (6·2) | 194,033 (9·8) | 604,868 (11·6) | 812,126 (18·7) | 98,922 (6·3) | 164,820 (12·4) |
| Redness | 283,345 (6·7) | 529,175 (14·5) | 66,134 (2·7) | 104,933 (5·3) | 295,413 (5·7) | 512,516 (11·8) | 58,375 (3·7) | 128,223 (9·6) |
| Itching | 299,407 (7·1) | 504,016 (13·8) | 72,095 (2·9) | 95,356 (4·8) | 309,607 (5·9) | 466,319 (10·7) | 66,469 (4·2) | 139,314 (10·5) |
| **Any systemic reaction** | 2,444,362 (57·9) | 2,752,592 (75·6) | 1,088,296 (43·8) | 1,226,561 (61·9) | 2,972,931 (57·1) | 3,237,621 (74·5) | 600,498 (38·4) | 781,299 (58·8) |
| Fatigue | 1,624,531 (38·5) | 2,221,361 (61·0) | 643,206 (25·9) | 904,556 (45·7) | 1,941,979 (37·3) | 2,588,541 (59·6) | 353,226 (22·6) | 569,758 (42·9) |
| Headache | 1,349,155 (31·9) | 1,906,337 (52·4) | 460,786 (18·6) | 690,138 (34·8) | 1,595,091 (30·6) | 2,226,046 (51·2) | 236,380 (15·1) | 397,675 (29·9) |
| Myalgia | 954,469 (22·6) | 1,724,474 (47·4) | 450,562 (18·1) | 726,994 (36·7) | 1,219,190 (23·4) | 2,085,722 (48·0) | 204,146 (13·0) | 392,448 (29·5) |
| Chills | 451,583 (10·7) | 1,202,364 (33·0) | 172,283 (6·9) | 459,577 (23·2) | 542,285 (10·4) | 1,426,710 (32·8) | 89,261 (5·7) | 253,475 (19·1) |
| Fever | 446,178 (10·6) | 1,182,201 (32·5) | 187,713 (7·6) | 478,912 (24·2) | 565,804 (10·9) | 1,449,504 (33·4) | 76,288 (4·9) | 230,073 (17·3) |
| Joint pain | 444,630 (10·5) | 1,023,525 (28·1) | 188,846 (7·6) | 400,963 (20·2) | 539,196 (10·3) | 1,214,624 (28·0) | 102,810 (6·6) | 226,303 (17·0) |
| Nausea | 447,766 (10·6) | 728,730 (20·0) | 106,872 (4·3) | 161,455 (8·2) | 500,782 (9·6) | 794,450 (18·3) | 61,491 (3·9) | 106,653 (8·0) |
| Diarrhea | 272,890 (6·5) | 313,252 (8·6) | 106,079 (4·3) | 101,107 (5·1) | 323,773 (6·2) | 352,077 (8·1) | 59,803 (3·8) | 66,967 (5·0) |
| Abdominal pain | 179,210 (4·2) | 283,422 (7·8) | 50,991 (2·1) | 71,115 (3·6) | 203,575 (3·9) | 316,165 (7·3) | 29,936 (1·9) | 42,942 (3·2) |
| Rash | 65,498 (1·6) | 79,092 (2·2) | 19,193 (0·8) | 19,735 (1·0) | 70,985 (1·4) | 79,913 (1·8) | 14,781 (0·9) | 19,965 (1·5) |
| Vomiting | 43,998 (1·0) | 75,650 (2·1) | 10,936 (0·4) | 14,915 (0·8) | 49,483 (0·9) | 81,733 (1·9) | 6,227 (0·4) | 9,994 (0·8) |

Data are n (%) unless otherwise stated.
\*Reports of local and systemic reactions are not mutually exclusive.

**Supplemental Table 6:** Most common local and systemic reactions^*^ to mRNA COVID-19 vaccine reported in v-safe, by dose and severity,^†^ 0-7 days after vaccination with BNT162b2 vaccine.

|  | Day | Dose 1 |  |  |  | Dose 2 |  |  |  |
| --- | --- | --- | --- | --- | --- | --- | --- | --- | --- |
|  |  | All, n | Severe | Moderate | Mild | All, n | Severe | Moderate | Mild |
| Injection site pain | 0 | 2,272,335 | 2,533 (0.1) | 80,358 (3.5) | 599,511 (26.4) | 1,766,510 | 4,359 (0.2) | 94,156 (5.3) | 503,779 (28.5) |
|  | 1 | 2,545,271 | 18,827 (0.7) | 334,755 (13.2) | 1,289,293 (50.7) | 2,027,330 | 48,810 (2.4) | 453,726 (22.4) | 885,434 (43.7) |
|  | 2 | 2,545,434 | 4,356 (0.2) | 62,838 (2.5) | 565,455 (22.2) | 2,116,614 | 10,391 (0.5) | 126,741 (6.0) | 680,408 (32.1) |
|  | 3 | 2,507,344 | 2,119 (0.1) | 26,602 (1.1) | 216,785 (8.6) | 2,067,908 | 3,332 (0.2) | 43,037 (2.1) | 336,222 (16.3) |
|  | 4 | 2,436,977 | 1,420 (0.1) | 16,710 (0.7) | 102,077 (4.2) | 2,028,926 | 1,820 (0.1) | 20,548 (1.0) | 149,408 (7.4) |
|  | 5 | 2,332,032 | 1,138 (0.05) | 11,965 (0.5) | 60,401 (2.6) | 2,000,426 | 1,272 (0.1) | 12,719 (0.6) | 73,486 (3.7) |
|  | 6 | 2,249,409 | 909 (0.04) | 8,901 (0.4) | 40,597 (1.8) | 2,000,472 | 1,106 (0.1) | 11,008 (0.6) | 46,707 (2.3) |
|  | 7 | 2,198,611 | 768 (0.03) | 7,783 (0.4) | 32,967 (1.5) | 2,067,201 | 1,557 (0.1) | 14,159 (0.7) | 44,322 (2.1) |
| Fatigue | 0 | 2,272,335 | 7,280 (0.3) | 79,232 (3.5) | 193,192 (8.5) | 1,766,510 | 11,293 (0.6) | 98,802 (5.6) | 185,051 (10.5) |
|  | 1 | 2,545,271 | 35,734 (1.4) | 229,606 (9.0) | 326,015 (12.8) | 2,027,330 | 135,581 (6.7) | 523,998 (25.8) | 373,601 (18.4) |
|  | 2 | 2,545,434 | 16,936 (0.7) | 114,562 (4.5) | 211,046 (8.3) | 2,116,614 | 39,668 (1.9) | 217,269 (10.3) | 308,126 (14.6) |
|  | 3 | 2,507,344 | 10,636 (0.4) | 74,341 (3.0) | 145,114 (5.8) | 2,067,908 | 15,361 (0.7) | 104,673 (5.1) | 193,174 (9.3) |
|  | 4 | 2,436,977 | 8,275 (0.3) | 58,170 (2.4) | 109,266 (4.5) | 2,028,926 | 10,280 (0.5) | 69,886 (3.4) | 133,603 (6.6) |
|  | 5 | 2,332,032 | 7,062 (0.3) | 49,739 (2.1) | 88,721 (3.8) | 2,000,426 | 8,089 (0.4) | 55,840 (2.8) | 103,919 (5.2) |
|  | 6 | 2,249,409 | 6,428 (0.3) | 44,044 (2.0) | 76,633 (3.4) | 2,000,472 | 7,200 (0.4) | 48,388 (2.4) | 86,907 (4.3) |
|  | 7 | 2,198,611 | 6,027 (0.3) | 40,428 (1.8) | 67,168 (3.1) | 2,067,201 | 7,528 (0.4) | 46,669 (2.3) | 78,361 (3.8) |
| Headache | 0 | 2,272,335 | 3,394 (0.1) | 42,501 (1.9) | 167,985 (7.4) | 1,766,510 | 5,217 (0.3) | 52,759 (3.0) | 144,892 (8.2) |
|  | 1 | 2,545,271 | 20,011 (0.8) | 129,629 (5.1) | 265,970 (10.4) | 2,027,330 | 82,393 (4.1) | 333,605 (16.5) | 381,368 (18.8) |
|  | 2 | 2,545,434 | 10,458 (0.4) | 69,347 (2.7) | 162,658 (6.4) | 2,116,614 | 24,063 (1.1) | 134,054 (6.3) | 249,895 (11.8) |
|  | 3 | 2,507,344 | 6,670 (0.3) | 46,850 (1.9) | 110,115 (4.4) | 2,067,908 | 10,356 (0.5) | 68,461 (3.3) | 148,990 (7.2) |
|  | 4 | 2,436,977 | 5,552 (0.2) | 38,319 (1.6) | 85,635 (3.5) | 2,028,926 | 7,238 (0.4) | 47,550 (2.3) | 103,204 (5.1) |
|  | 5 | 2,332,032 | 4,911 (0.2) | 34,379 (1.5) | 72,831 (3.1) | 2,000,426 | 6,154 (0.3) | 40,322 (2.0) | 82,191 (4.1) |
|  | 6 | 2,249,409 | 4,733 (0.2) | 31,540 (1.4) | 64,890 (2.9) | 2,000,472 | 5,467 (0.3) | 35,177 (1.8) | 69,168 (3.5) |
|  | 7 | 2,198,611 | 4,381 (0.2) | 29,475 (1.3) | 58,752 (2.7) | 2,067,201 | 5,372 (0.3) | 34,057 (1.6) | 63,628 (3.1) |
| Myalgia | 0 | 2,272,335 | 1,999 (0.1) | 29,601 (1.3) | 96,095 (4.2) | 1,766,510 | 4,001 (0.2) | 38,960 (2.2) | 75,790 (4.3) |
|  | 1 | 2,545,271 | 18,440 (0.7) | 136,939 (5.4) | 219,125 (8.6) | 2,027,330 | 101,801 (5.0) | 408,637 (20.2) | 293,241 (14.5) |
|  | 2 | 2,545,434 | 7,441 (0.3) | 56,954 (2.2) | 112,788 (4.4) | 2,116,614 | 23,521 (1.1) | 140,700 (6.6) | 209,074 (9.9) |
|  | 3 | 2,507,344 | 4,200 (0.2) | 33,605 (1.3) | 65,696 (2.6) | 2,067,908 | 6,925 (0.3) | 54,206 (2.6) | 100,982 (4.9) |
|  | 4 | 2,436,977 | 3,255 (0.1) | 25,814 (1.1) | 46,369 (1.9) | 2,028,926 | 4,146 (0.2) | 32,786 (1.6) | 60,489 (3.0) |
|  | 5 | 2,332,032 | 2,831 (0.1) | 22,598 (1.0) | 37,598 (1.6) | 2,000,426 | 3,239 (0.2) | 25,326 (1.3) | 44,242 (2.2) |
|  | 6 | 2,249,409 | 2,543 (0.1) | 20,904 (0.9) | 33,016 (1.5) | 2,000,472 | 2,973 (0.1) | 22,422 (1.1) | 36,522 (1.8) |
|  | 7 | 2,198,611 | 2,504 (0.1) | 19,474 (0.9) | 30,222 (1.4) | 2,067,201 | 3,379 (0.2) | 23,046 (1.1) | 33,563 (1.6) |
| Chills | 0 | 2,272,335 | 879 (0.04) | 8,246 (0.4) | 34,000 (1.5) | 1,766,510 | 2,091 (0.1) | 14,428 (0.8) | 38,195 (2.2) |
|  | 1 | 2,545,271 | 8,558 (0.3) | 45,518 (1.8) | 78,033 (3.1) | 2,027,330 | 62,884 (3.1) | 210,579 (10.4) | 207,218 (10.2) |
|  | 2 | 2,545,434 | 3,371 (0.1) | 18,659 (0.7) | 36,412 (1.4) | 2,116,614 | 11,744 (0.6) | 51,490 (2.4) | 76,276 (3.6) |
|  | 3 | 2,507,344 | 1,462 (0.1) | 9,241 (0.4) | 19,569 (0.8) | 2,067,908 | 2,582 (0.1) | 13,423 (0.6) | 25,421 (1.2) |
|  | 4 | 2,436,977 | 1,051 (0.04) | 6,915 (0.3) | 13,967 (0.6) | 2,028,926 | 1,336 (0.1) | 7,424 (0.4) | 14,223 (0.7) |
|  | 5 | 2,332,032 | 863 (0.04) | 5,531 (0.2) | 11,284 (0.5) | 2,000,426 | 955 (0.05) | 5,423 (0.3) | 10,583 (0.5) |
|  | 6 | 2,249,409 | 779 (0.03) | 5,048 (0.2) | 9,932 (0.4) | 2,000,472 | 851 (0.04) | 4,763 (0.2) | 9,029 (0.5) |
|  | 7 | 2,198,611 | 752 (0.03) | 4,645 (0.2) | 8,889 (0.4) | 2,067,201 | 1,222 (0.1) | 5,515 (0.3) | 9,039 (0.4) |
| Joint pain | 0 | 2,272,335 | 1,069 (0.05) | 11,375 (0.5) | 24,689 (1.1) | 1,766,510 | 2,396 (0.1) | 18,677 (1.1) | 25,699 (1.5) |
|  | 1 | 2,545,271 | 9,676 (0.4) | 61,691 (2.4) | 69,532 (2.7) | 2,027,330 | 55,446 (2.7) | 225,949 (11.2) | 137,601 (6.8) |
|  | 2 | 2,545,434 | 4,608 (0.2) | 31,238 (1.2) | 44,072 (1.7) | 2,116,614 | 14,386 (0.7) | 80,490 (3.8) | 90,461 (4.3) |
|  | 3 | 2,507,344 | 2,675 (0.1) | 19,912 (0.8) | 29,313 (1.2) | 2,067,908 | 4,624 (0.2) | 32,971 (1.6) | 46,467 (2.2) |
|  | 4 | 2,436,977 | 2,165 (0.1) | 15,923 (0.7) | 22,386 (0.9) | 2,028,926 | 2,882 (0.1) | 20,861 (1.0) | 29,916 (1.5) |
|  | 5 | 2,332,032 | 1,999 (0.1) | 13,922 (0.6) | 18,869 (0.8) | 2,000,426 | 2,341 (0.1) | 16,528 (0.8) | 23,366 (1.2) |
|  | 6 | 2,249,409 | 1,773 (0.1) | 13,018 (0.6) | 16,874 (0.8) | 2,000,472 | 2,138 (0.1) | 15,046 (0.8) | 19,649 (1.0) |
|  | 7 | 2,198,611 | 1,686 (0.1) | 12,245 (0.6) | 15,605 (0.7) | 2,067,201 | 2,462 (0.1) | 15,782 (0.8) | 18,678 (0.9) |

Data are n (%) unless otherwise stated.
\*Top five reactions determined by reported frequency after dose 2 of both mRNA COVID-19 vaccines in v-safe, excluding fever because it was not rated mild/moderate/severe. Symptoms are not mutually exclusive. †Mild was defined as “noticeable symptoms but they aren’t a problem”, moderate was defined as “symptoms that limit normal activities, and severe symptoms”, and severe symptoms “make normal daily activities difficult or impossible”.

**Supplemental Table 7:**
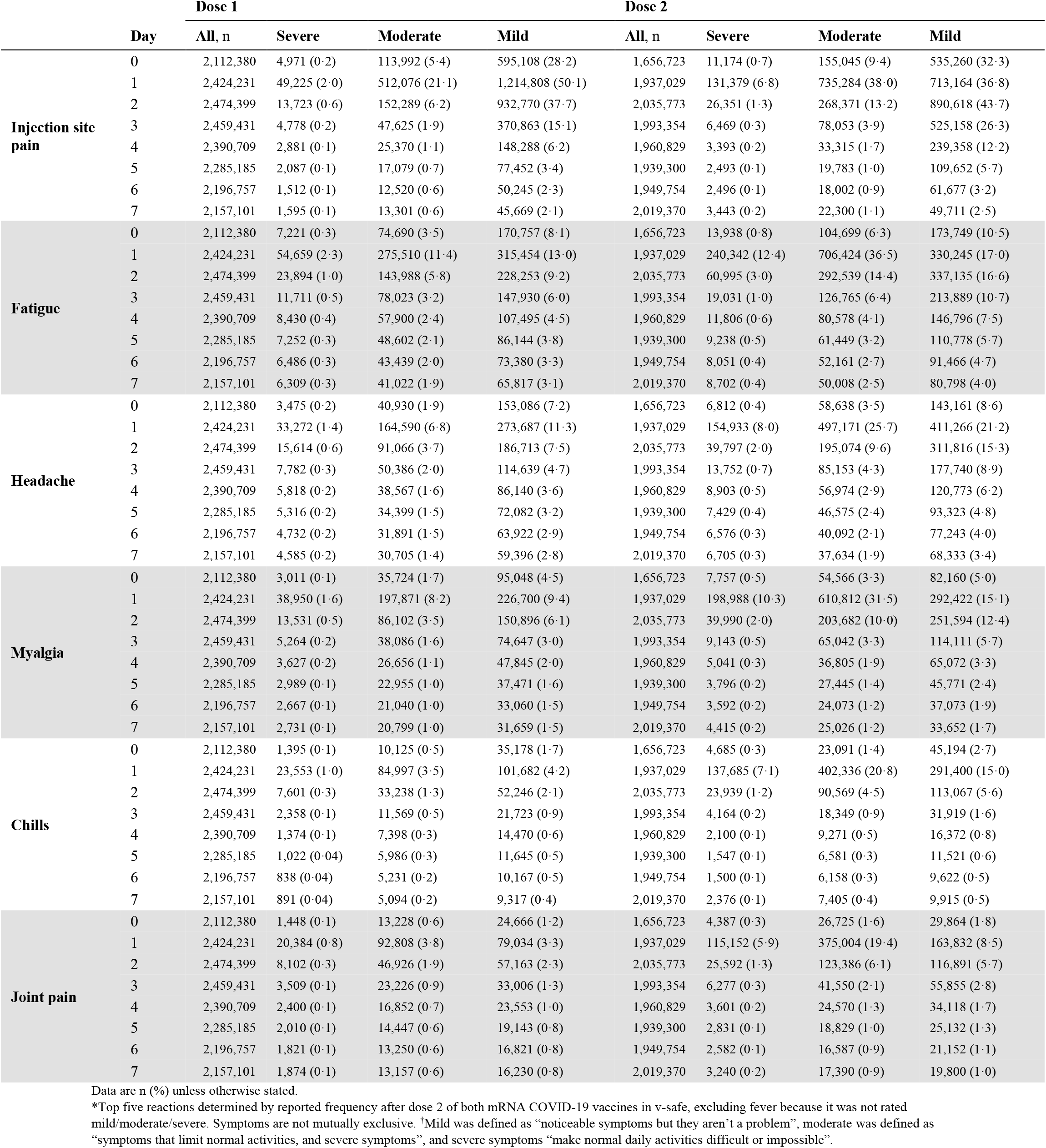
Most common local and systemic reactions^*^ to mRNA COVID-19 vaccine reported in v-safe, by dose and severity,^†^ 0-7 days after vaccination with mRNA-1273 vaccine.

**Supplemental Table 8:**
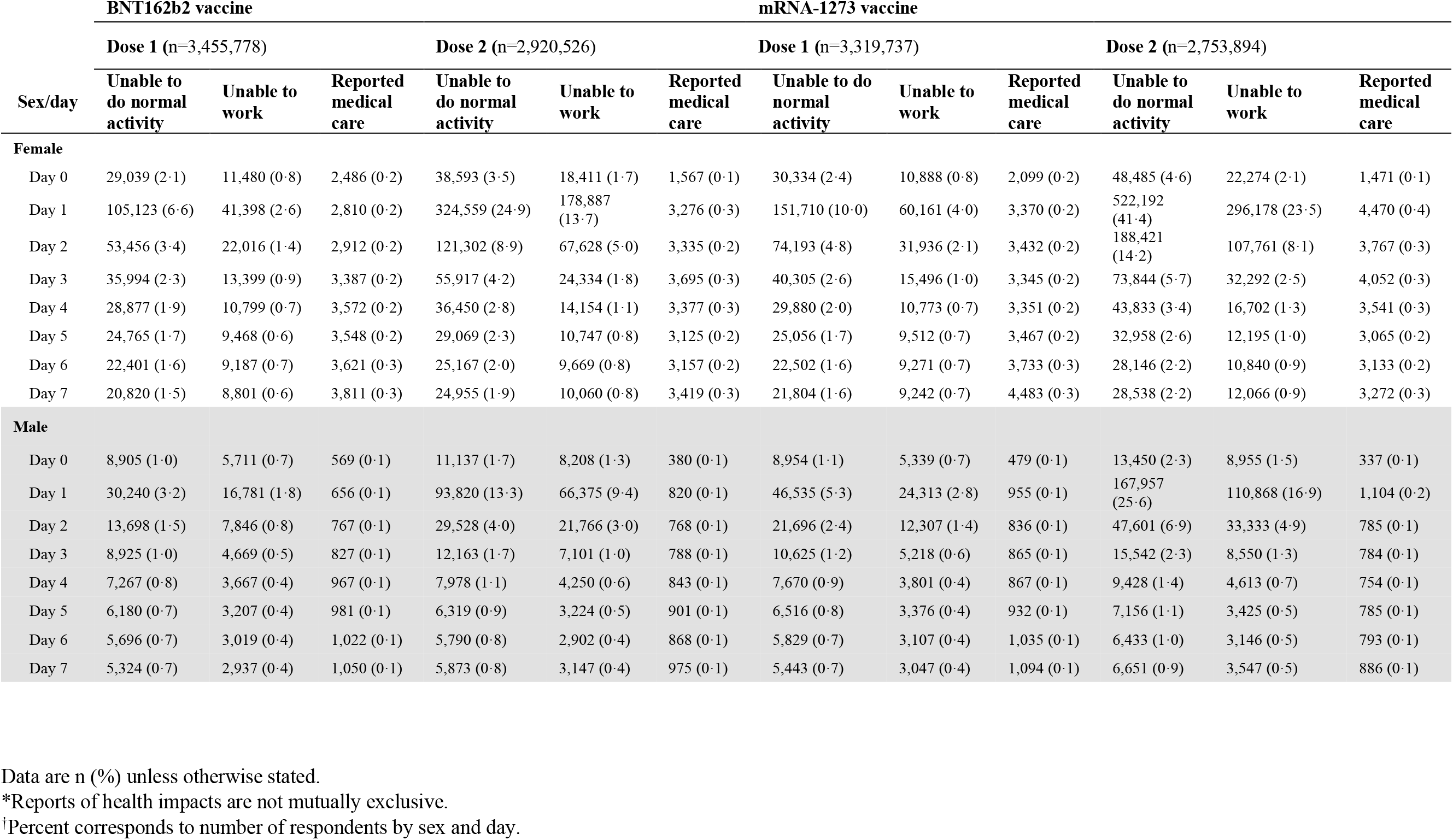
Reported health impact^*^ 0-7 days after vaccination by mRNA COVID-19 vaccine manufacturer, dose, and sex reported in v-safe—December 14, 2020–June 14, 2021.

**Supplemental Figure 1:**
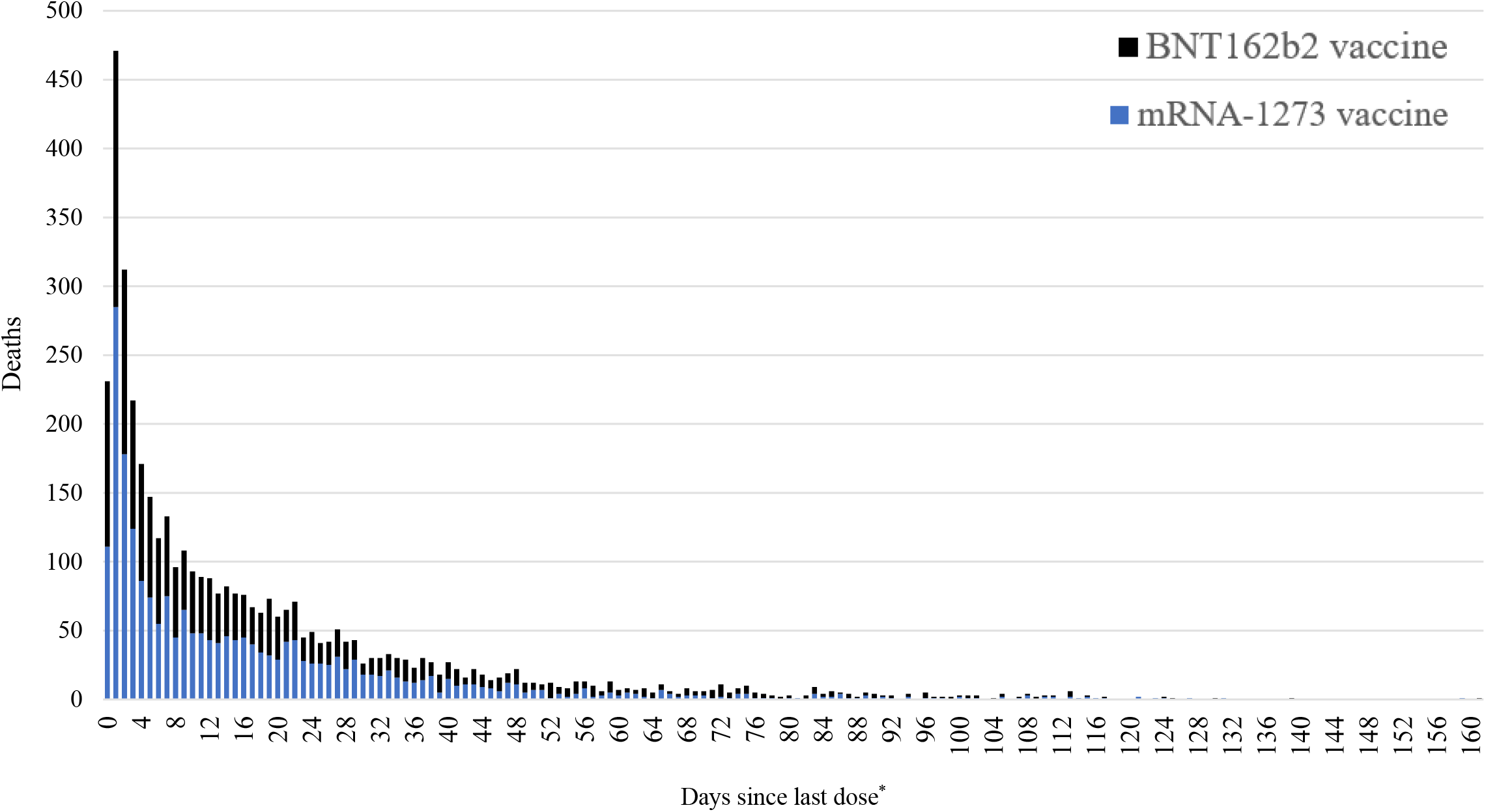
Number of reports of death per day following vaccination, by manufacturer, to Vaccine Adverse Event Reporting System (VAERS)—December 14, 2020–June 14, 2021. ^*^x-axis reports through 161 days since last dose.

**Supplemental Figure 2:**
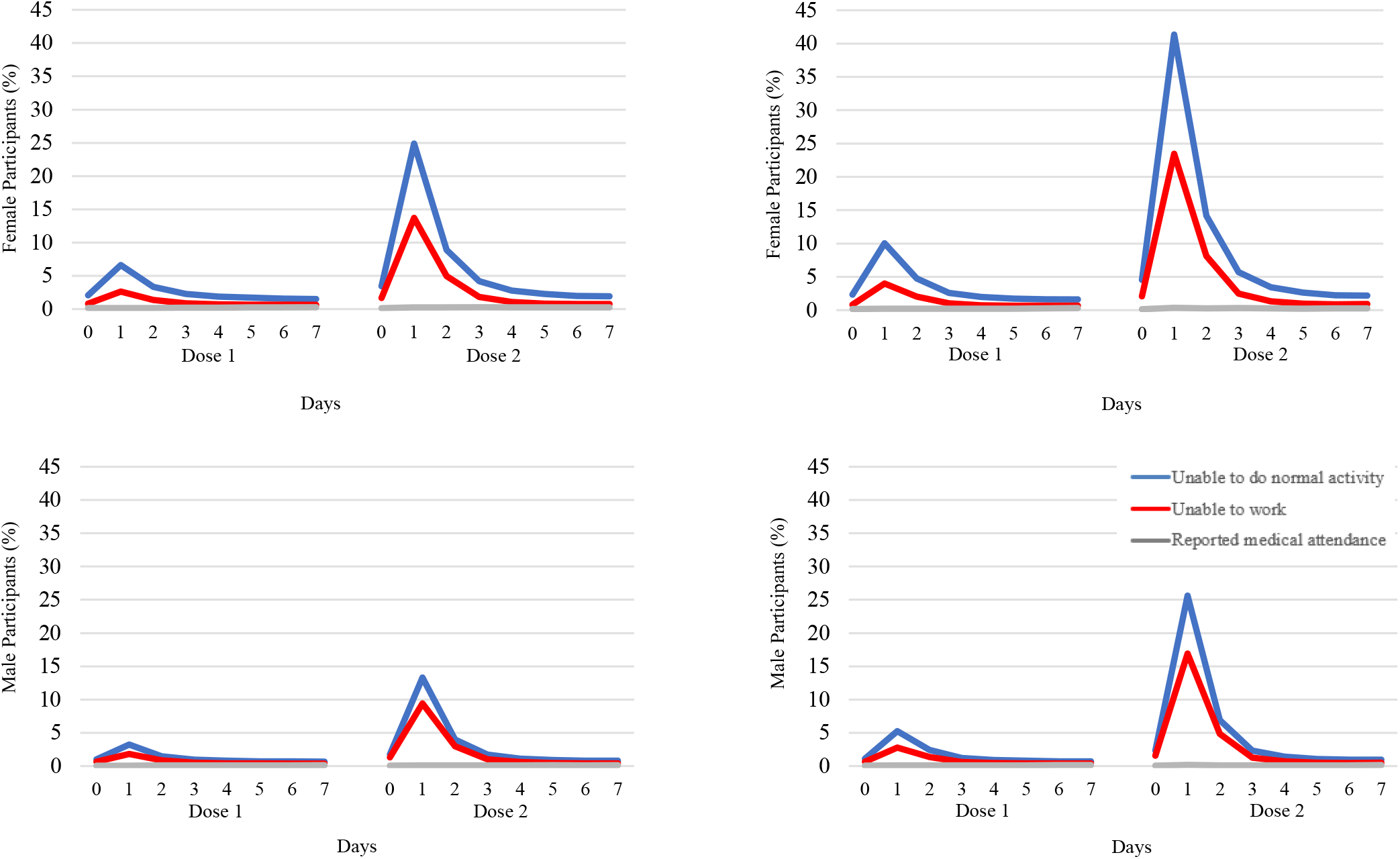
Reported health impact 0-7 days after mRNA COVID-19 vaccination by manufacturer, type of impact, and sex reported in v-safe—December 14, 2020–June 14, 2021. *Top left:* Female participants reporting health impact after receiving BNT162b2 vaccine. *Top right:* Female participants reporting health impact after receiving mRNA-1273 vaccine. *Bottom left:* Male participants reporting health impact after receiving BNT162b2 vaccine. *Bottom right*: Male participants reporting health impact after receiving mRNA-1273 vaccine.

## References

1. Oliver SE, Gargano JW, Marin M, et al. The Advisory Committee on Immunization Practices’ Interim Recommendation for Use of Pfizer-BioNTech COVID-19 Vaccine - United States, December 2020. MMWR Morbidity and mortality weekly report 2020; 69(50): 1922–4.

2. Oliver SE, Gargano JW, Marin M, et al. The Advisory Committee on Immunization Practices’ Interim Recommendation for Use of Moderna COVID-19 Vaccine - United States, December 2020. MMWR Morbidity and mortality weekly report 2021; 69(5152): 1653–6.

3. Polack FP, Thomas SJ, Kitchin N, et al. Safety and Efficacy of the BNT162b2 mRNA Covid-19 Vaccine. The New England Journal of Medicine 2020; 383(27): 2603–15.

4. Baden LR, El Sahly HM, Essink B, et al. Efficacy and Safety of the mRNA-1273 SARS-CoV-2 Vaccine. The New England Journal of Medicine 2021; 384(5): 403–16.

5. Anderson EJ, Rouphael NG, Widge AT, et al. Safety and Immunogenicity of SARS-CoV-2 mRNA-1273 Vaccine in Older Adults. New England Journal of Medicine 2020; 383(25): 2427–38.

6. Chen RT, Davis RL, Rhodes PH. Special methodological issues in pharmacoepidemiology studies of vaccine safety. In: Strom BL, editor. Pharmacoepidemiology. 4th John Wiley & Sons; Sussex: 2005

7. Dooling K, McClung N, Chamberland M, et al. The Advisory Committee on Immunization Practices’ Interim Recommendation for Allocating Initial Supplies of COVID-19 Vaccine - United States, 2020. MMWR Morbidity and mortality weekly report 2020; 69(49): 1857–9.

8. Shimabukuro TT, Nguyen M, Martin D, DeStefano F. Safety monitoring in the Vaccine Adverse Event Reporting System (VAERS). Vaccine 2015; 33(36): 4398–405.

9. https://www.cdc.gov/coronavirus/2019-ncov/vaccines/safety/vsafe.html

10. Gee J, Marquez P, Su J, et al. First Month of COVID-19 Vaccine Safety Monitoring - United States, December 14, 2020-January 13, 2021. MMWR Morbidity and mortality weekly report 2021; 70(8): 283–8.

11. Chapin-Bardales J, Gee J, Myers T. Reactogenicity Following Receipt of mRNA-Based COVID-19 Vaccines. JAMA 2021; 325(21): 2201–2.

12. Shimabukuro TT, Cole M, Su JR. Reports of Anaphylaxis After Receipt of mRNA COVID-19 Vaccines in the US—December 14, 2020-January 18, 2021. JAMA 2021; 325(11): 1101–2.

13. Shay DK, Gee J, Su JR, et al. Safety Monitoring of the Janssen (Johnson & Johnson) COVID-19 Vaccine - United States, March-April 2021. MMWR Morbidity and mortality weekly report 2021; 70(18): 680–4.

14. Allergic Reactions Including Anaphylaxis After Receipt of the First Dose of Pfizer-BioNTech COVID-19 Vaccine - United States, December 14-23, 2020. MMWR Morbidity and mortality weekly report 2021; 70(2): 46–51.

15. https://www.cdc.gov/vaccinesafety/ensuringsafety/monitoring/vaers/index.html

16. Code of Federal Regulations Title 21 https://www.accessdata.fda.gov/scripts/cdrh/cfdocs/cfcfr/cfrsearch.cfm?fr

17. https://www.cdc.gov/vaccinesafety/pdf/VAERS-v2-SOP.pdf

18. https://www.cdc.gov/nchs/nvss/leading-causes-of-death.htm

19. https://covid.cdc.gov/covid-data-tracker/#datatracker-home

20. Abara WE, Gee J, Mu Y, et al. Expected Rates of Select Adverse Events following Immunization for COVID-19 Vaccine Safety Monitoring. medRxiv 2021: 2021.08.31.21262919.

21. Szarfman A, Machado SG, O’Neill RT. Use of screening algorithms and computer systems to efficiently signal higher-than-expected combinations of drugs and events in the US FDA’s spontaneous reports database. Drug Saf 2002; 25(6): 381–92.

22. Martin, D, Menschik D, Bryant-Genevier M, Ball R. Data mining for prospective early detection of safety signals in the Vaccine Adverse Event Reporting System (VAERS): a case study of febrile seizures after a 2010-2011 seasonal influenza virus vaccine. Drug Saf. 2013; Jul:36(7):547-56.

23. https://www.cdc.gov/vaccinesafety/ensuringsafety/monitoring/vsd/index.html

24. https://www.cdc.gov/vaccines/acip/work-groups-vast/index.html

25. https://www.cdc.gov/vaccines/acip/meetings/index.html

26. MacNeil JR, Su JR, Broder KR, et al. Updated Recommendations from the Advisory Committee on Immunization Practices for Use of the Janssen (Johnson & Johnson) COVID-19 Vaccine After Reports of Thrombosis with Thrombocytopenia Syndrome Among Vaccine Recipients - United States, April 2021. MMWR Morbidity and mortality weekly report 2021; 70(17): 651–6.

27. Gargano JW, Wallace M, Hadler SC, et al. Use of mRNA COVID-19 Vaccine After Reports of Myocarditis Among Vaccine Recipients: Update from the Advisory Committee on Immunization Practices - United States, June 2021. MMWR Morbidity and mortality weekly report 2021; 70(27): 977–82.

28. Rosenblum HG, Hadler SC, Moulia D et al. Use of COVID-19 Vaccines After Reports of Adverse Events Among Adult Recipients of Janssen (Johnson & Johnson) and mRNA COVID-19 Vaccines (Pfizer-BioNTech and Moderna): Update from the Advisory Committee on Immunization Practices – United States, July 2021. MMWR Morbidity and mortality weekly report 2021: 70(32); 1094-1099.

29. Menni C, Klaser K, May A, et al. Vaccine side-effects and SARS-CoV-2 infection after vaccination in users of the COVID Symptom Study app in the UK: a prospective observational study. Lancet Infect Dis. 2021; 21(7):939-949

30. Hervé C, Laupèze B, Del Giudice G, Didierlaurent AM, Tavares Da Silva F. The how’s and what’s of vaccine reactogenicity. NPJ Vaccines 2019;4: 39.

31. Zimmermann P, Curtis N. Factors That Influence the Immune Response to Vaccination. Clin Microbiol Rev 2019; 32(2).

32. Klein SL, Jedlicka A, Pekosz A. The Xs and Y of immune responses to viral vaccines. Lancet Infect Dis 2010; 10(5): 338–49.

33. Pathiravasan CH, Zhang Y, Trinquart L, et al. Adherence of Mobile App-Based Surveys and Comparison With Traditional Surveys: eCohort Study. J Med Internet Res 2021; 23(1): e24773.

34. Guo X, Vittinghoff E, Olgin JE, Marcus GM, Pletcher MJ. Volunteer Participation in the Health eHeart Study: A Comparison with the US Population. Sci Rep 2017; 7(1): 1956.

35. Millar MM, Elena JW, Gallicchio L, et al. The feasibility of web surveys for obtaining patient-reported outcomes from cancer survivors: a randomized experiment comparing survey modes and brochure enclosures. BMC Med Res Methodol 2019; 19(1): 208.

36. Messer BL, Dillman DA. Surveying the General Public over the Internet Using Address-Based Sampling and Mail Contact Procedures. Public Opinion Quarterly 2011; 75(3): 429–57.

37. https://www.kff.org/coronavirus-covid-19/poll-finding/kff-covid-19-vaccine-monitor-april-2021/

38. https://www.kff.org/coronavirus-covid-19/poll-finding/kff-covid-19-vaccine-monitor-june-2021/

39. https://www.whitehouse.gov/briefing-room/statements-releases/2021/04/21/fact-sheet-president-biden-to-call-on-all-employers-to-provide-paid-time-off-for-employees-to-get-vaccinated-after-meeting-goal-of-200-million-shots-in-the-first-100-days/

40. Miller ER, Lewis P, Shimabukuro TT, et al. Post-licensure safety surveillance of zoster vaccine live (Zostavax®) in the United States, Vaccine Adverse Event Reporting System (VAERS), 2006-2015. Human vaccines & immunotherapeutics 2018; 14(8): 1963–9

41. Moro PL, Haber P, McNeil MM. Challenges in evaluating post-licensure vaccine safety: observations from the Centers for Disease Control and Prevention. Expert Review of Vaccines 2019; 18(10): 1091–101

42. Haber P, Moro PL, Ng C, et al. Safety review of tetanus toxoid, reduced diphtheria toxoid, acellular pertussis vaccines (Tdap) in adults aged ≥65 years, Vaccine Adverse Event reporting System (VAERS), United States, September 2010-December 2018. Vaccine 2020; 38(6): 1476–80

43. Woo EJ, Moro PL. Postmarketing safety surveillance of quadrivalent recombinant influenza vaccine: Reports to the vaccine adverse event reporting system. Vaccine 2021; 39(13): 1812–7.

44. https://www.cdc.gov/vaccines/covid-19/clinical-considerations/managing-anaphylaxis.html?CDC_AA_refVal=https%3A%2F%2Fwww.cdc.gov%2Fvaccines%2Fcovid-19%2Finfo-by-product%2Fpfizer%2Fanaphylaxis-management.html

45. Abu Mouch S, Roguin A, Hellou E, et al. Myocarditis following COVID-19 mRNA vaccination. Vaccine 2021.

46. Marshall M, Ferguson ID, Lewis P, et al. Symptomatic Acute Myocarditis in Seven Adolescents Following Pfizer-BioNTech COVID-19 Vaccination. Pediatrics 2021 Jun 4;e2021052478. Online ahead of print

47. https://www.cdc.gov/vaccines/covid-19/clinical-considerations/myocarditis.html

48. Moro PL, Arana J, Cano M, Lewis P, Shimabukuro TT. Deaths Reported to the Vaccine Adverse Event Reporting System, United States, 1997-2013. Clin Infect Dis 2015; 61(6): 980–7.

49. https://wonder.cdc.gov/vaers.html

50. Murphy SL, Xu J, Kochanek KD, Arias E, Tejada-Vera B. Deaths: Final Data for 2018. Natl Vital Stat Rep 2021; 69(13): 1–83.

51. https://www.cdc.gov/vaccinesafety/pdf/SCK_COVID_Vaccine_Mortality-508.pdf

52. https://www.cdc.gov/coronavirus/2019-ncov/vaccines/distributing/first-doses.html

53. https://vaers.hhs.gov/data/dataguide.html

54. https://www.cdc.gov/coronavirus/2019-ncov/science/science-briefs/fully-vaccinated-people.html

